# The skin we live in: pigmentation traits and tanning behaviour in British young adults, an observational and genetically-informed study

**DOI:** 10.1101/2022.01.08.22268938

**Authors:** Carolina Bonilla, Cilia Mejia-Lancheros

**Author notes:** Corresponding author Carolina Bonilla Departamento de Medicina Preventiva Faculdade de Medicina Universidade de São Paulo Av. Dr. Arnaldo 455 CEP 01246-903 São Paulo Brazil.

## Abstract

**Background:** Skin cancer incidence has been increasing worldwide, representing a particularly high burden for populations of European ancestry. Outdoor and indoor tanning using ultraviolet radiation (UVR) devices are major risk factors for skin cancer. While tanning behaviours can be modified by targeted interventions to reduce skin cancer rates, there is insufficient evidence on the motivations for tanning preferences and their relationship with pigmentation phenotypes. The present observational and genetically-informed study investigates motives for tanning and the role that pigmentation phenotypes play on outdoor and indoor tanning behaviour in British young adults.

**Methods:** This study included 3722 participants from the Avon Longitudinal Study of Parents and Children in South West England. Skin, hair and eye colour features, and tanning ability and preferences were collected using a questionnaire applied when participants were ∼25 years of age. Genotypes for 41 single nucleotide polymorphisms (SNPs) associated with pigmentation were obtained from a subset of participants who provided a biological sample, and used to estimate the probability of having particular pigmentation traits with the HIrisPlex-S system.

**Results:** Liking to tan and outdoor tanning were strongly influenced by skin, hair and eye pigmentation, and tanning ability. However, the association of these traits with UV indoor tanning was weaker. Conversely, females, participants of lower socioeconomic position, individuals who were unhappy with their pigmentation phenotype during adolescence, and participants who believed that tanning helps prevent sunburn were more likely to have used UVR-based tanning devices.

**Conclusion:** Our results provide evidence to support the implementation of skin cancer preventative interventions that consider individual biological characteristics and motives for undergoing outdoor and indoor tanning.

## Introduction

The incidence of melanoma and non-melanoma skin cancer (NMSC), which comprises basal cell carcinoma (BCC) and squamous cell carcinoma (SCC), has been steadily increasing in all populations for the past three decades. The Global Burden of Disease study reported that between 1990 and 2017, SCC experienced an increase of 310%, BCC increased by 77%, while melanoma increased by 161%^1^. In particular, populations of European descent are greatly affected by skin cancer^2^. In 2018 melanoma age-standardised (world) incidence rate was 33.6 in Australia, 15.0 in the UK and 12.4 in Canada, compared to 2.8 in Brazil, 2.2 in Mexico and 0.6 in Nigeria^3^.

Ultraviolet radiation (UVR), with wavelengths between 100 and 400 nm, is the leading risk factor for skin cancer. Intermittent exposure and sun burning, especially during childhood has been associated with melanoma, whereas chronic, cumulative exposure more often results in NMSC onset^4^. Whilst most of UVR exposure occurs through natural sunlight, artificial or indoor tanning, defined as the use of an UVR emission device to produce a cosmetic tan^5^, represents an important UVR source. Indoor tanning is quite frequent in high-income countries, predominantly among young people^6^. For example, in 2010 the prevalence in England of having ever used a sunbed was ∼6% amongst 11 to 17-year olds^7^. The prevalence in Scotland and Wales for the same age range was ∼14% and 11%, respectively. In addition, a comprehensive systematic review and meta-analysis of indoor tanning in the United States, northern and western Europe, and Australia, that assessed literature reports until 2013, found an overall prevalence of 36% in adults, 55% in university students, and 19% in adolescents^8^. More recent studies suggest that these rates are declining, although they remain high^9,10,11^.

Indoor tanning has been associated with an increased risk of melanoma and NMSC, and thus, was classified as a group I carcinogen by the International Agency for Research on Cancer (IARC) in 2009^12^. A systematic review and meta-analysis of 12 studies of indoor tanning and NMSC showed that ever exposure to indoor tanning was associated with a higher risk of both, BCC (RR 1.29; 95% CI 1.08, 1.53) and SCC (RR 1.67; 95%CI 1.29, 2.17)^5^. Similarly, melanoma risk was higher due to sunbed use ever, as reported by a systematic review and meta-analysis of 27 observational studies (RR 1.20; 95% CI 1.08, 1.34)^13^. Exposure at a young age (< 25 years old) increased these risks considerably^5, 13^.

On the other hand, the synthesis of vitamin D in the skin is highly dependent on UVR. Vitamin D underlies bone and muscle health, and its deficiency has been associated with risk of a number of complex diseases such as multiple sclerosis, type 2 diabetes, Alzheimer’s disease, and breast and colorectal cancer^14^. Beyond the production of vitamin D there are other UVR-related benefits including inhibition of autoimmune reactivity and reduction of blood pressure via the generation of nitric oxide^14–16^. Consequently, achieving a balance between producing adequate levels of vitamin D while limiting the damage to the skin has become an active area of debate in public health^17, 18^.

In order to implement interventions that curve tanning it is crucial to understand the motivations and personal characteristics (cultural, behavioural and biological) behind individual UVR exposure. Several studies have investigated risk factors associated with indoor tanning, consistently finding that female sex, younger age, pigmentation traits and appearance enhancement were strong predictors for it^6, 19–21^. A few have considered attitudes towards tanning, with a focus on college students, and usually framing the question with respect to the importance of tanning for the respondent 9,22,^23^. However, none has explored the potentially causal association between pigmentation phenotypes and tanning behaviour and motives using genetic data.

This study investigated the determinants of “liking to tan”, as well as the reasons given for this choice. We examined the association of pigmentation traits, reported via questionnaire and predicted using genetic variants, with tanning preferences, in a cohort of young adults from the South West of England, who are part of the Avon Longitudinal Study of Parents and Children (ALSPAC). Additionally, we assessed the relationship between tanning and participants’ willingness to change their skin and hair colour, and the reaction their skin colour elicited in other people.

There are many well-known and potential predictors of tanning behaviour that are worth examining in this population, more so since it is possible for researchers to assess the consequences of that behaviour on the next generation of ALSPAC children (ALSPAC-G2), who are being recruited to the cohort as the first generation (G1) completes their families^24^. In this article we emphasised pigmentation characteristics as drivers for tanning, with the understanding that non-pigmentation-related risk factors are likely to play a role as well and their consideration in future studies is warranted.

## Materials and methods

### Study design

We performed an observational analysis in the first generation (G1) of the ALSPAC birth cohort using mostly answers to one of its latest questionnaires, and a genetically-informed analysis applying the HIrisPlex-S system^15^ to estimate probabilities of pigmentation phenotypes. Participants included in the study were of White ethnicity.

### Study population

The ALSPAC cohort recruited pregnant women resident in Avon, UK with expected dates of delivery between 1st April 1991 to 31st December 1992. The initial number of pregnancies enrolled was 14,541 (for these at least one questionnaire has been returned or a “Children in Focus” clinic has been attended by 19/07/99). Of these initial pregnancies, there was a total of 14,676 foetuses, resulting in 14,062 live births and 13,988 children who were alive at 1 year of age. When the oldest children were approximately 7 years of age, an attempt was made to bolster the initial sample with eligible cases who had failed to join the study originally. As a result, when considering variables collected from the age of seven onwards (and potentially abstracted from obstetric notes) there are data available for more than the 14,541 pregnancies mentioned above. The number of new pregnancies not in the initial sample (known as Phase I enrolment) that are currently represented on the built files and reflecting enrolment status at the age of 24 is 913 (456, 262 and 195 recruited during Phases II, III and IV respectively), resulting in an additional 913 children being enrolled. The phases of enrolment are described in more detail in the cohort profile paper and its update. The total sample size for analyses using any data collected after the age of seven is therefore 15,454 pregnancies, resulting in 15,589 foetuses. Of these 14,901 were alive at 1 year of age^25–27^. A 10% sample of the ALSPAC cohort, known as the Children in Focus (CiF) group, attended clinics at the University of Bristol at various time intervals between 4 to 61 months of age. The CiF group were chosen at random from the last 6 months of ALSPAC births (1432 families attended at least one clinic). Excluded were those mothers who had moved out of the area or were lost to follow-up, and those partaking in another study of infant development in Avon.

Study data were collected and managed using Research Electronic Data Capture (REDCap) tools hosted at the University of Bristol (England)^28, 29^. REDCap is a secure, web-based software platform designed to support data capture for research studies.

Please note that the study website contains details of all the data that is available through a fully searchable data dictionary and variable search tool (http://www.bristol.ac.uk/alspac/researchers/our-data/).

Ethical approval for the study was obtained from the ALSPAC Ethics and Law Committee and the Local Research Ethics Committees (http://www.bristol.ac.uk/alspac/researchers/research-ethics/). Consent for biological samples has been collected in accordance with the Human Tissue Act (2004). Informed consent for the use of data collected via questionnaires and clinics was obtained from participants following the recommendations of the ALSPAC Ethics and Law Committee at the time.

### Exposures and outcomes

We used data extracted from the Life@25+ questionnaire, Tanning and Sun Exposure section, which was applied to ALSPAC young people (YP) in 2017-2018, when they were ∼25 years old. The questionnaire mainly inquired about outdoor and indoor tanning preferences, reasons for engaging in any type of tanning, sun exposure and sun protection habits, and pigmentation traits. Participants were also asked about skin cancer history in the family and their knowledge about the relationship between tanning and skin cancer. Life@25+ replicated some of the questions about sun exposure and pigmentation traits that were asked in previous ALSPAC questionnaires^30^. A few of the earlier responses were used here to compare childhood parent-reported phenotypes and experiences with those in adulthood conveyed by the Life@25+ survey.

Variables related to whether YP were trying to change their skin or hair colour to match media images, having been hurt or name-called because of their skin colour, seeing someone being bullied because of that person’s skin colour, and the teachers’ equal treatment of pupils regardless of skin colour, were obtained when YP were between 12 and 14 years of age. Information about parental education (< O level, O level and > O level) and socioeconomic position (SEP) (manual and non-manual occupation) were taken from prior ALSPAC’s questionnaires as well.

### Genotypes

ALSPAC YP were genotyped using the Illumina HumanHap550 quad chip (Illumina, Inc., San Diego, CA) by the Wellcome Trust Sanger Institute, Cambridge, UK and the Laboratory Corporation of America, Burlington, NC, US. The resulting raw genome-wide data were subjected to standard quality control methods. Individuals were excluded on the basis of sex mismatches; minimal (<0.325) or excessive heterozygosity (>0.345); disproportionate levels of individual missingness (>3%), cryptic relatedness measured as proportion of identity by descent (IBD > 0.1) and insufficient sample replication (IBD < 0.8). The remaining individuals were assessed for evidence of population stratification by multidimensional scaling analysis and compared with Hapmap II (release 22) European descent (CEU), Han Chinese (CHB), Japanese (JPT) and Yoruba (YRI) reference populations; all individuals with non-European ancestry were removed. Single nucleotide polymorphisms (SNPs) with a minor allele frequency of < 1%, a call rate of < 95% or evidence for violations of Hardy-Weinberg equilibrium (p<5x10^-7^) were removed. Genotypic data were subsequently imputed using Markov Chain Haplotyping software and phased haplotype data from the Thousand Genomes Project (2010-11 data freeze) that included 1092 samples of mixed ethnicity who had had singleton and monomorphic sites removed.

We applied the HIrisPlex-S system (https://hirisplex.erasmusmc.nl/) to 8564 individuals with available genotypes, to calculate the probability of them showing each of 14 characteristics (blue, intermediate or brown eyes; brown, blond, red, or black hair; light or dark hair; very pale, pale, intermediate, dark and dark to black skin)^31^. HIrisPlex-S was developed in European populations for forensic research and uses genotypes of 41 SNPs in pigmentation genes, of which we had 39 SNPs available. Variants rs312262906 (currently rs796296176) and rs201326893 in *MC1R* were missing in all participants, whereas a few others were missing at random in some participants. We used the SNP rs116927526, in strong linkage disequilibrium (LD) with rs312262906 (r^2^ = 1) in the British population, as a replacement. Rs201326893 is monomorphic in the British population according to LDlink (https://ldlink.nci.nih.gov/), and therefore we were not able to use a proxy SNP to replace this variant. Missing genotypes affect prediction performance, which was assessed with the area under the curve (AUC) measure. Full AUC values and mean AUC loss for each trait are shown in Supplementary Table 1. Typically, HIrisPlex-S is employed in forensics to predict these pigmentation traits as part of the identification of a DNA sample donor. In this study, instead of actual predicted phenotypes, we used their prediction probabilities as independent variables in the regression models.

### Statistical analysis

Binary and ordered logistic and linear regressions were used in the observational and the genetically-informed analyses. Adjustment for age and sex was applied in the observational analysis, while age, sex, and the top 10 genetic principal components (PCs) were adjusted for in the genetically-informed analysis.

We ran pairwise correlation tests to assess the relationship of pigmentation and sun exposure traits reported in childhood with those informed in the Life@25+ questionnaire, and the relationship of HIrisPlex-S probabilities with the top 10 genetic PCs. Based on these results we tested some of the models controlling for the top 5 rather than the top 10 PCs as a sensitivity analysis.

All analyses were performed using the Stata software package v16 and 17.

### Text-based data analysis

Text answers provided as an explanation to the “other” option in questions related to hair colour, eye colour, reasons for tanning, and sun protective actions, were recoded to create inclusive categories based on specific themes and explore interdependence (coincidences) among these variables. We used the Stata commands *txttool* and *precoin* (part of the user-written command called *coin*), to investigate coincidences with network analysis^32^.

Six, 8, 17 and 9 events (categories) were created for hair colour, eye colour, reason for tanning and sun protective measures, respectively. Each category was defined by precise key terms and included at least two responses. Responses that did not fit into any pre-established category were grouped into an “other” category. Answers that mentioned two or more key terms were allocated to all of the corresponding categories. The connections of the 17 reason-for-tanning categories were graphically explored in a multidimensional network plot, whilst the relationship between sun-protection categories was examined using a dendrogram (coincidence cluster). Both graphs were generated with the *coin* command in Stata^32^.

## Results

### Description of the characteristics of the study population

The present study included 3722 White ALSPAC YP who responded to the Tanning and Sun Exposure section of the Life@25+ questionnaire, 2825 of whom had genotypic data available.

The characteristics of the study population are shown in Table 1.

**Table 1.** Characteristics of ALSPAC young people who completed the questionnaire Life@25+, Tanning and Sun Exposure section.

|  |  |  |  |
| --- | --- | --- | --- |
| **YP age at questionnaire completion (years)** | **mean** | **SD** | **N** |
|  | 25.3 | 0.6 | 3722 |
| **YP sex** | **n** | **% male** | **N** |
| male | 1271 | 34.2 | 3722 |
| female | 2451 | 75.8 |  |
| **mother's social class<sup>a</sup>** | **n** | **%** | **N** |
| manual | 424 | 13.7 | 3100 |
| non-manual | 2676 | 86.3 |  |
| **father's social class<sup>a</sup>** | **n** | **%** | **N** |
| manual | 1163 | 35.3 | 3295 |
| non-manual | 2132 | 64.7 |  |
| **mother's education<sup>a</sup>** | **n** | **%** | **N** |
| < O level | 652 | 18.5 | 3520 |
| O level | 1189 | 33.8 |  |
| > O level | 1679 | 47.7 |  |
| **YP likes to tan** | **yes/no/missing** | **% yes** | **N** |
|  | 1921/1790/11 | 51.6 | 3722 |
| **how?** | **yes/no** | **% yes** | **N<sup>b</sup>** |
| usually tans outdoors | 1731/198 | 89.7 | 1929 |
| usually tans indoors (sun bed/sun lamp/tanning booth) | 309/1621 | 16.0 | 1930 |
| usually tans indoors (spray tan) | 172/1758 | 8.9 | 1930 |
| usually tans indoors (self-tanning lotions or creams) | 522/1408 | 27.1 | 1930 |
| **why?** | **yes/no** | **% yes** | **N<sup>b</sup>** |
| it gives YP more confidence | 1272/658 | 65.9 | 1930 |
| it makes YP feel happier | 1377/550 | 71.5 | 1927 |
| makes YP look better in photos | 1094/836 | 56.7 | 1930 |
| makes YP look thinner | 486/1444 | 25.2 | 1930 |
| conceals body imperfections | 541/1389 | 28.0 | 1930 |
| YP looks more attractive to others | 940/990 | 48.7 | 1930 |
| pale skin is unattractive | 423/1507 | 21.9 | 1930 |
| protects YP from the sun | 100/1830 | 5.2 | 1930 |
| another reason | 175/1755 | 9.1 | 1930 |
| **skin colour** | **n** | **%** | **N** |
| very fair | 755 | 20.3 | 3714 |
| fair | 2205 | 59.4 |  |
| olive | 647 | 17.4 |  |
| brown | 107 | 2.9 |  |
| **eye colour** | **n** | **%** | **N** |
| blue | 1613 | 43.4 | 3715 |
| green | 774 | 20.8 |  |
| grey | 130 | 3.5 |  |
| brown | 1017 | 27.4 |  |
| other | 181 | 4.9 |  |
| **natural hair colour at 18 years old** | **n** | **%** | **N** |
| red | 151 | 4.1 | 3718 |
| blonde | 693 | 18.6 |  |
| light brown | 1379 | 37.1 |  |
| dark brown | 1388 | 37.3 |  |
| black | 60 | 1.6 |  |
| other | 47 | 1.3 |  |
| **YP has freckles** | **n** | **%** | **N** |
| no | 1579 | 42.5 | 3716 |
| yes, a few | 1628 | 43.8 |  |
| yes, many | 509 | 13.7 |  |
| **skin colour changes after being in the sun (tanning ability)** | **n** | **%** | **N** |
| always burns never tans | 232 | 6.3 | 3705 |
| burns easily rarely tans | 1272 | 34.3 |  |
| doesn't change | 193 | 5.2 |  |
| tans easily rarely burns | 1752 | 47.3 |  |
| always tans never burns | 115 | 3.1 |  |
| can't say, skin always protected | 141 | 3.8 |  |
| **YP had a red/painful sunburn in the past 2 years lasting a day or more** | **n** | **%** | **N** |
| never | 806 | 22.6 | 3572 |
| once | 997 | 27.9 |  |
| twice | 972 | 27.2 |  |
| 3 times | 426 | 11.9 |  |
| 4 times | 228 | 6.4 |  |
| 5 times or more | 143 | 4.0 |  |
| **sun protection** | **yes/no** | **% yes** | **N** |
| YP does not use any protection whilst out in the sun | 244/3473 | 6.6 | 3717 |
| YP wears a hat whilst out in the sun | 1066/2651 | 28.7 |  |
| YP wears clothing whilst out in the sun | 1366/2351 | 36.8 |  |
| YP wears sunblock/sunscreen whilst out in the sun | 3439/278 | 92.5 |  |
| YP avoids the sun | 784/2933 | 21.1 |  |
| YP uses other methods of protection | 34/3683 | 0.9 |  |
| **sunblock/sunscreen factor** | **n** | **%** | **N** |
| < 15 | 166 | 4.8 | 3433 |
| 15-24 | 1056 | 30.8 |  |
| 25-49 | 1528 | 44.5 |  |
| 50 or higher | 683 | 19.9 |  |
| **frequency YP applies sunblock/sunscreen whilst out in the sun** | **n** | **%** | **N** |
| once only | 603 | 17.5 | 3435 |
| every 3-4 hours | 1883 | 54.8 |  |
| every 2 hours | 741 | 21.6 |  |
| every hour | 189 | 5.5 |  |
| every half hour | 19 | 0.6 |  |
| **use of indoor tanning equipment such as sun bed/sun lamp/tanning booth** |  |  |  |
| YP has ever used indoor tanning equipment | **yes/no** | **% yes** | **N** |
|  | 942/2774 | 25.4 | 3716 |
| age YP first started using indoor tanning equipment | **mean** | **SD** | **N** |
|  | 19.2 | 3.1 | 926 |
| use of indoor tanning equipment in the past 12 months | **n** | **%** | **N** |
| not used | 487 | 51.5 | 946<sup>b</sup> |
| once or twice a year | 141 | 14.9 |  |
| a few times in the year | 199 | 21.0 |  |
| once a month | 59 | 6.2 |  |
| once a week | 44 | 4.7 |  |
| more than once a week | 16 | 1.7 |  |
| **skin cancer** | **yes/no** | **% yes** | **N** |
| YP has been diagnosed with skin cancer | 9/3707 | 0.2 | 3716 |
| YP has family member diagnosed with skin cancer | 448/3266 | 12.1 | 3714 |
| **beliefs about tanning** | **yes/no/don't know** | **% yes** | **N** |
| YP believes indoor tanning helps prevent sunburn | 284/2624/808 | 7.6 | 3716 |
| YP thinks indoor tanning using sun bed/sun lamp can cause skin cancer | 3164/108/441 | 85.2 | 3713 |
| **body image<sup>a</sup>** | **yes/no** | **% yes** | **N** |
| YP is trying to change skin colour | 269/2691 | 9.1 | 2960 |
| YP is trying to change hair colour | 197/2763 | 6.7 | 2960 |
| YP has been hurt because of their skin colour | 34/3076 | 1.1 | 3110 |
| YP has been called names because of their skin colour | 76/3032 | 2.5 | 3108 |
| YP has seen someone bullied because of their skin colour | 910/2189 | 29.4 | 3099 |
| YP's teachers treat everybody the same, regardless of skin colour<br>(0=agree/1=disagree) | 120/2748 | 4.2 | 2868 |

<sup>a</sup>Variables were obtained from questionnaires applied at earlier stages of the cohort.
<sup>b</sup>Some YP with a missing answer for the first question provided an answer for a following question derived from the former.

**Table 2.**
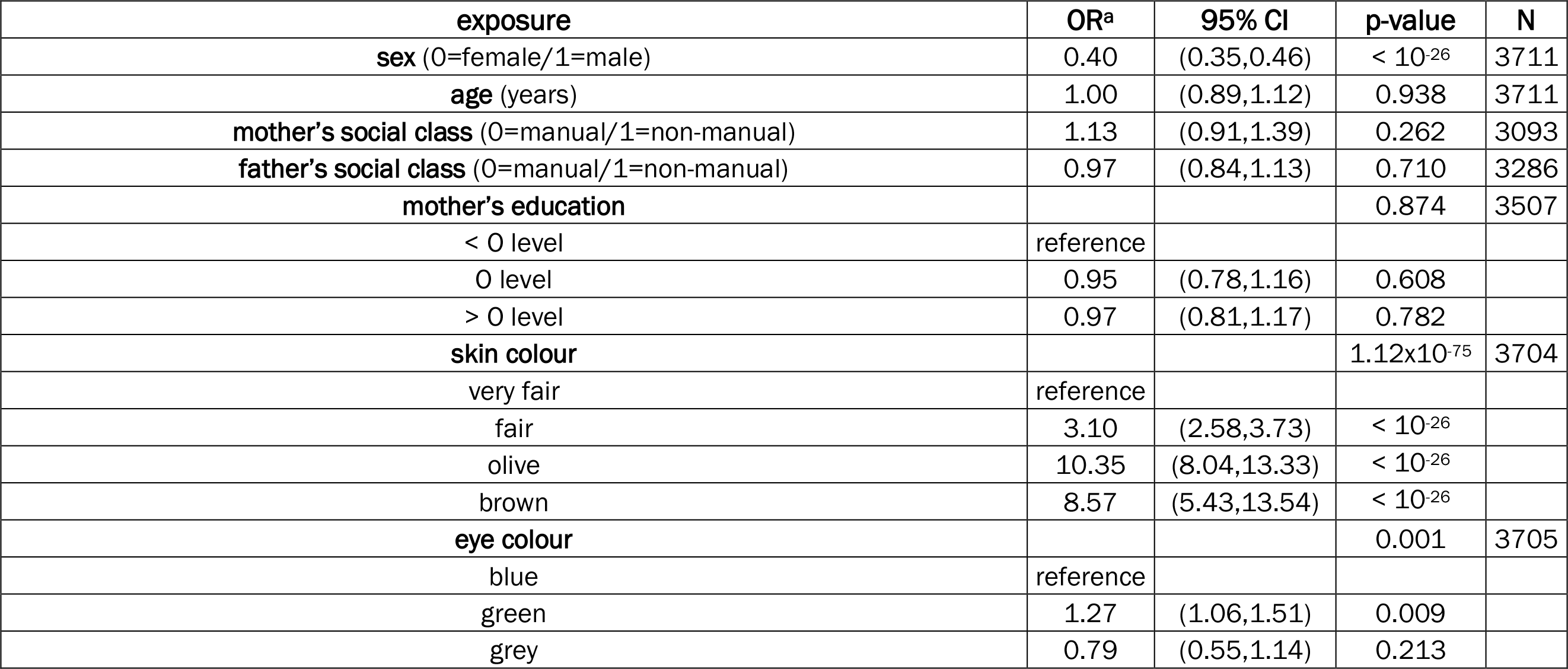
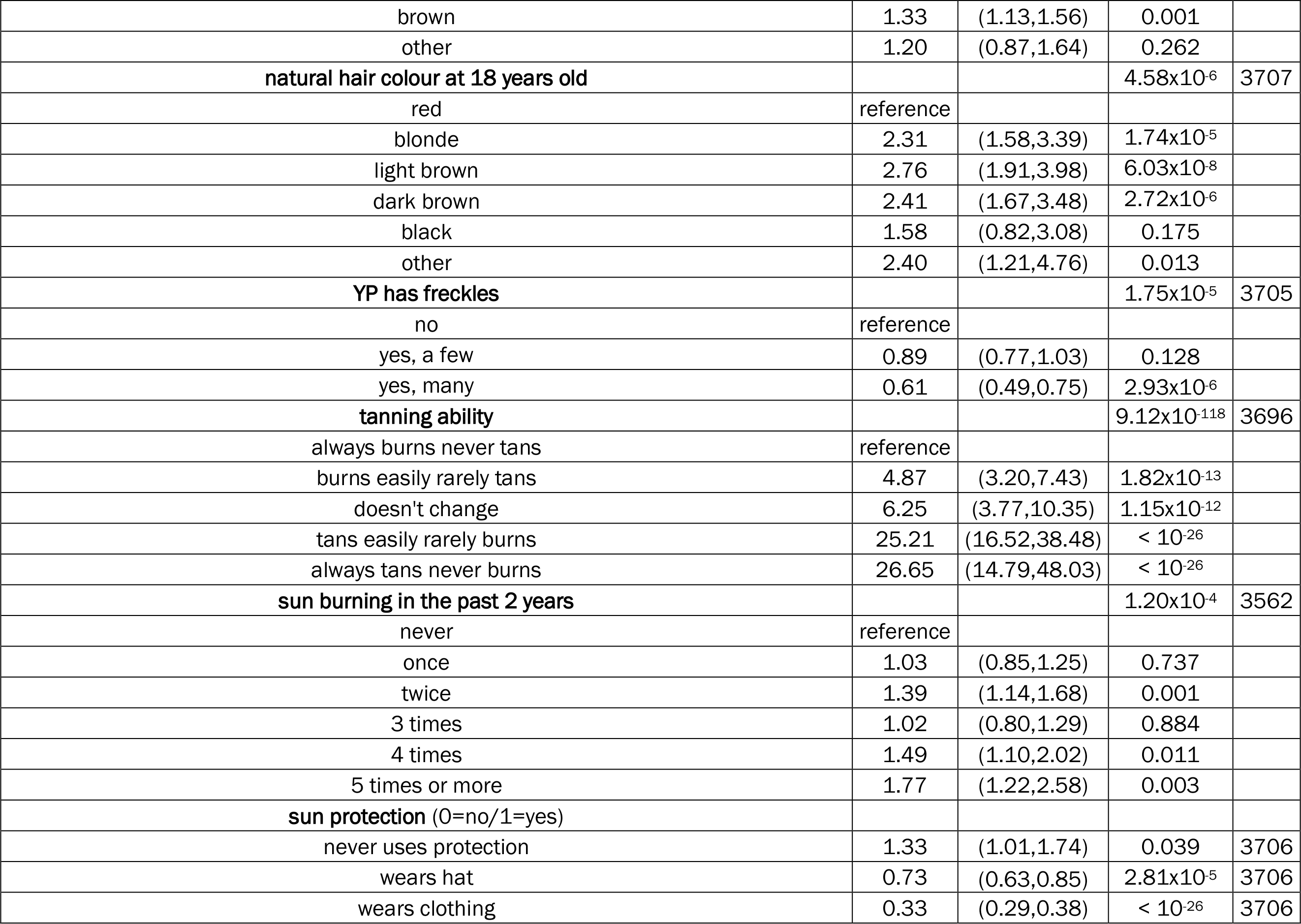
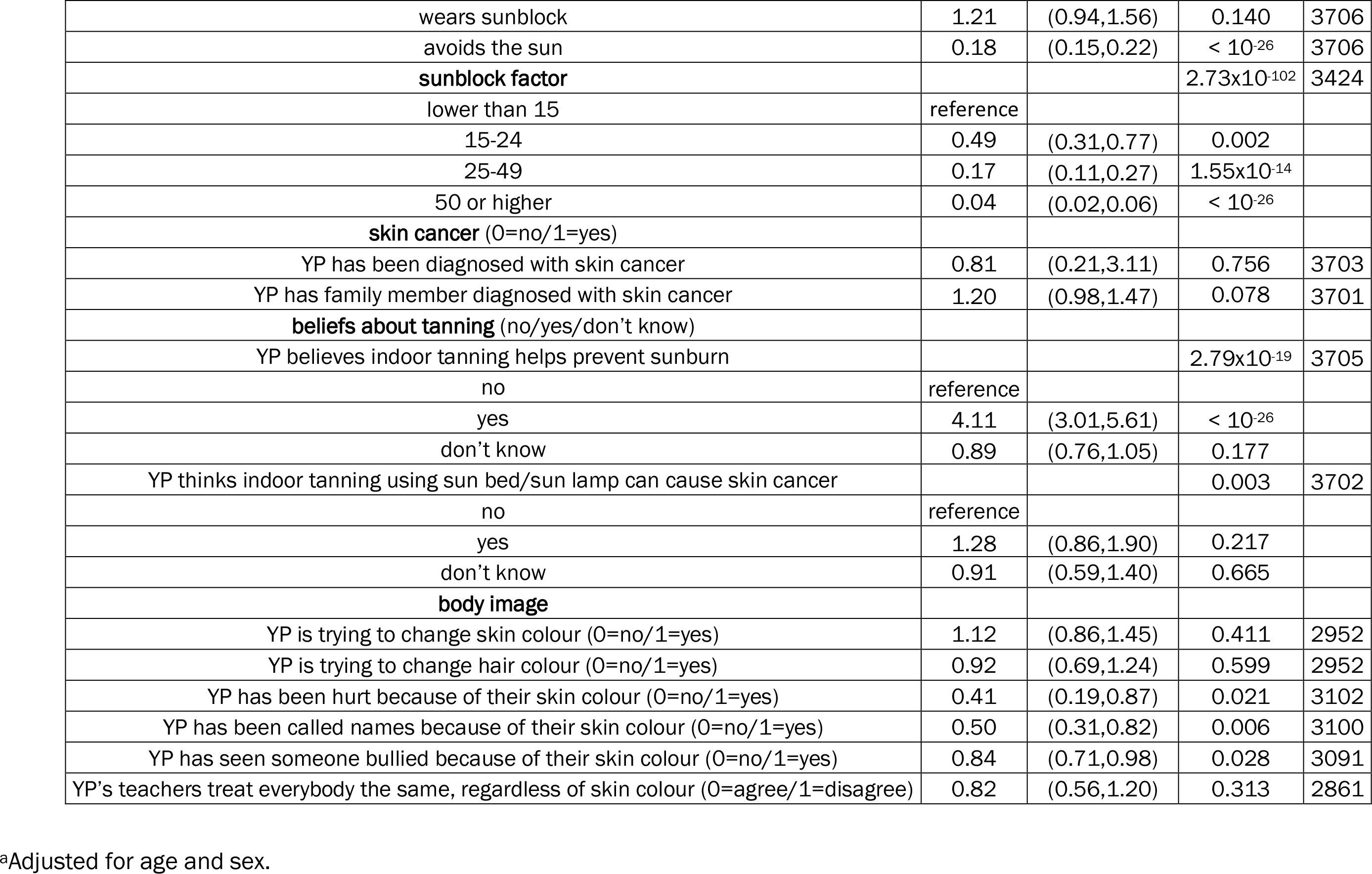
Association of socio-demographic, pigmentation-related and sun exposure factors with ‘liking to tan’ in ALSPAC young people.

| **exposure** | **OR<sup>a</sup>** | **95% CI** | **p-value** | **N** |
| --- | --- | --- | --- | --- |
| **sex** (0=female/1=male) | 0.40 | (0.35,0.46) | < 10<sup>-26</sup> | 3711 |
| **age** (years) | 1.00 | (0.89,1.12) | 0.938 | 3711 |
| **mother's social class** (0=manual/1=non-manual) | 1.13 | (0.91,1.39) | 0.262 | 3093 |
| **father's social class** (0=manual/1=non-manual) | 0.97 | (0.84,1.13) | 0.710 | 3286 |
| **mother's education** |  |  | 0.874 | 3507 |
| < 0 level | reference |  |  |  |
| 0 level | 0.95 | (0.78,1.16) | 0.608 |  |
| > 0 level | 0.97 | (0.81,1.17) | 0.782 |  |
| **skin colour** |  |  | 1.12x10<sup>-75</sup> | 3704 |
| very fair | reference |  |  |  |
| fair | 3.10 | (2.58,3.73) | < 10<sup>-26</sup> |  |
| olive | 10.35 | (8.04,13.33) | < 10<sup>-26</sup> |  |
| brown | 8.57 | (5.43,13.54) | < 10<sup>-26</sup> |  |
| **eye colour** |  |  | 0.001 | 3705 |
| blue | reference |  |  |  |
| green | 1.27 | (1.06,1.51) | 0.009 |  |
| grey | 0.79 | (0.55,1.14) | 0.213 |  |
| brown | 1.33 | (1.13,1.56) | 0.001 |  |
| other | 1.20 | (0.87,1.64) | 0.262 |  |
| **natural hair colour at 18 years old** |  |  | 4.58x10<sup>-6</sup> | 3707 |
| red | reference |  |  |  |
| blonde | 2.31 | (1.58,3.39) | 1.74x10<sup>-5</sup> |  |
| light brown | 2.76 | (1.91,3.98) | 6.03x10<sup>-8</sup> |  |
| dark brown | 2.41 | (1.67,3.48) | 2.72x10<sup>-6</sup> |  |
| black | 1.58 | (0.82,3.08) | 0.175 |  |
| other | 2.40 | (1.21,4.76) | 0.013 |  |
| **YP has freckles** |  |  | 1.75x10<sup>-5</sup> | 3705 |
| no | reference |  |  |  |
| yes, a few | 0.89 | (0.77,1.03) | 0.128 |  |
| yes, many | 0.61 | (0.49,0.75) | 2.93x10<sup>-6</sup> |  |
| **tanning ability** |  |  | 9.12x10<sup>-118</sup> | 3696 |
| always burns never tans | reference |  |  |  |
| burns easily rarely tans | 4.87 | (3.20,7.43) | 1.82x10<sup>-13</sup> |  |
| doesn't change | 6.25 | (3.77,10.35) | 1.15x10<sup>-12</sup> |  |
| tans easily rarely burns | 25.21 | (16.52,38.48) | < 10<sup>-26</sup> |  |
| always tans never burns | 26.65 | (14.79,48.03) | < 10<sup>-26</sup> |  |
| **sun burning in the past 2 years** |  |  | 1.20x10<sup>-4</sup> | 3562 |
| never | reference |  |  |  |
| once | 1.03 | (0.85,1.25) | 0.737 |  |
| twice | 1.39 | (1.14,1.68) | 0.001 |  |
| 3 times | 1.02 | (0.80,1.29) | 0.884 |  |
| 4 times | 1.49 | (1.10,2.02) | 0.011 |  |
| 5 times or more | 1.77 | (1.22,2.58) | 0.003 |  |
| **sun protection (0=no/1=yes)** |  |  |  |  |
| never uses protection | 1.33 | (1.01,1.74) | 0.039 | 3706 |
| wears hat | 0.73 | (0.63,0.85) | 2.81x10<sup>-5</sup> | 3706 |
| wears clothing | 0.33 | (0.29,0.38) | < 10<sup>-26</sup> | 3706 |
| wears sunblock | 1.21 | (0.94,1.56) | 0.140 | 3706 |
| avoids the sun | 0.18 | (0.15,0.22) | < 10<sup>-26</sup> | 3706 |
| **sunblock factor** |  |  | 2.73x10<sup>-102</sup> | 3424 |
| lower than 15 | reference |  |  |  |
| 15-24 | 0.49 | (0.31,0.77) | 0.002 |  |
| 25-49 | 0.17 | (0.11,0.27) | 1.55x10<sup>-14</sup> |  |
| 50 or higher | 0.04 | (0.02,0.06) | < 10<sup>-26</sup> |  |
| **skin cancer** (0=no/1=yes) |  |  |  |  |
| YP has been diagnosed with skin cancer | 0.81 | (0.21,3.11) | 0.756 | 3703 |
| YP has family member diagnosed with skin cancer | 1.20 | (0.98,1.47) | 0.078 | 3701 |
| **beliefs about tanning** (no/yes/don't know) |  |  |  |  |
| YP believes indoor tanning helps prevent sunburn |  |  | 2.79x10<sup>-19</sup> | 3705 |
| no | reference |  |  |  |
| yes | 4.11 | (3.01,5.61) | < 10<sup>-26</sup> |  |
| don't know | 0.89 | (0.76,1.05) | 0.177 |  |
| YP thinks indoor tanning using sun bed/sun lamp can cause skin cancer |  |  | 0.003 | 3702 |
| no | reference |  |  |  |
| yes | 1.28 | (0.86,1.90) | 0.217 |  |
| don't know | 0.91 | (0.59,1.40) | 0.665 |  |
| **body image** |  |  |  |  |
| YP is trying to change skin colour (0=no/1=yes) | 1.12 | (0.86,1.45) | 0.411 | 2952 |
| YP is trying to change hair colour (0=no/1=yes) | 0.92 | (0.69,1.24) | 0.599 | 2952 |
| YP has been hurt because of their skin colour (0=no/1=yes) | 0.41 | (0.19,0.87) | 0.021 | 3102 |
| YP has been called names because of their skin colour (0=no/1=yes) | 0.50 | (0.31,0.82) | 0.006 | 3100 |
| YP has seen someone bullied because of their skin colour (0=no/1=yes) | 0.84 | (0.71,0.98) | 0.028 | 3091 |
| YP's teachers treat everybody the same, regardless of skin colour (0=agree/1=disagree) | 0.82 | (0.56,1.20) | 0.313 | 2861 |

<sup>a</sup>Adjusted for age and sex.

Participants were on average 25.3 years old, female (75.8%) and had parents with high SEP (i.e. non-manual occupation and higher educational attainment). Slightly over half of the respondents declared they liked to tan, and most of them (∼90%) usually tanned outdoors. Individuals who usually tanned indoors as well represented 27.4% of the sample, including those who used a sunbed, sun lamp or tanning booth (16%), a spray tan (9%), and/or self-tanning lotions (27%).

The main reasons for liking to tan were feeling happier, more confident, looking better in photos, and looking more attractive to others (∼50% or more respondents selected these options). Other reasons given by over 20% of participants were concealing body imperfections, looking thinner and believing that pale skin is unattractive. Five percent claimed protection from the sun as a reason for liking to tan, whereas 9% wrote down another reason. Among these other reasons given as a text answer (Supplementary Table 2), the most frequent were those related to enjoying being in the sun (14.8%), liking tanned skin (14.5%), looking healthier (14.2%) and improving skin conditions (13.3%). Increasing vitamin D levels was stated in 5.2% of text answers. The relationship between these categories is shown in Supplementary Figure 1. Generally, participants provided more than one reason for liking to tan (coincidences). For example, participants who said that they “like tanned skin” were more likely to answer that tanning helps with skin problems, makes them have a glowing skin, look attractive, and feel warm/summery, and also answered that there were other reasons for their preference. The strongest relationship observed was that between “like/enjoy being in the sun” and “feels relaxing” (p < 0.001).

With respect to pigmentation traits, respondents were primarily fair, had blue eyes, brown hair, and a few or no freckles (Table 1). The frequencies of trait combinations are shown in Supplementary Figures 2-4. Females exhibited higher odds of having blond hair and light brown hair than males (OR 1.70; 95% CI 1.41, 2.05; OR 1.21, 95% CI 1.05, 1.40, respectively), whereas the opposite was true for brown or black hair (OR 0.60; 95% CI 0.52, 0.69). Categories of text answers created for hair and eye colour are shown in Supplementary Tables 3 and 4. Hazel eyes was the most common answer to the latter, probably because it was not provided as an option in the original question.

Most participants reported tanning easily and rarely burning, followed by burning easily and rarely tanning (Table 1). There was a strong correlation between pigmentation, tanning and sun exposure phenotypes conveyed at 25 years old and those gathered in earlier ALSPAC questionnaires (when YP were between 1.3 and 5.8 years old) (Supplementary Table 5).

Overall, YP practiced sun protection quite thoroughly, ∼93% mentioned using sunblock and a substantial fraction (∼64%) applied sunblock with a sun protection factor (SPF) of 25 or more (Table 1). This was a noteworthy increase with respect to the 38% of respondents whose parents reported using sunblock with SPF of 25 or more on them when they were ∼69 months old. Many participants wear a hat or clothing when out in the sun (29-37%). Red or painful sunburn lasting a day or more was experienced once or twice in the past two years in just over a half of subjects (Table 1). Among the most frequent text answers for other strategies of sun protection were “sitting in the shade” (27.1%) and “wearing sunglasses” (12.5%) (Supplementary Table 6). Supplementary Figure 5 shows the hierarchical relationship of coincidences between sun protection measures. The closest relationship was observed between “using sunscreen products” and “other means” of protection. This relationship was in turn linked to wearing sunglasses, and the resulting relationship was then linked to sitting in the shade.

Asked about employing indoor tanning equipment such as a sunbed, sun lamp or tanning booth, 25.4% of YP manifested ever using it, and almost half of those individuals had done so in the past 12 months. Average age at first use was relatively late, at 19.2 (SD 3.1) years old, with men starting later than women (20.3 vs 19.1 years old, respectively, p = 0.0001). Intensive use of a sunbed, i.e. once a week or more, was carried out by a minority (6.4%) (Table 1).

Nine (0.2%) YP had been diagnosed with skin cancer, and 448 (12.1%) participants indicated having a close family member affected by the disease. Whilst over 85% of individuals were aware that UV indoor tanning can cause skin cancer, a few of them believed that it helps prevent sunburn (7.6%). However, there is a considerable number of participants who declare not knowing one way or the other (∼12% in the cancer question and ∼22% in the sun burning question) (Table 1).

### Observational analysis

#### 1. Liking to tan

The age and sex-adjusted associations between socio-demographic, pigmentation-related and sun exposure factors with “liking to tan” are presented in Table 2.

Female sex, darker skin colour, eye colour other than blue or grey, hair colour other than red, absence of freckles, tanning rather than burning, experiencing a painful sunburn several times in the past 2 years, and believing that indoor tanning helps prevent sunburn, were strong and statistically positive predictors of “liking to tan” (Table 2). Avoiding the sun and wearing clothing or a hat when out in the sun were inversely associated with tanning predilection. Individuals who “liked to tan” were more likely to apply sunblock than those who did not (OR 1.21; 95% CI 0.94, 1.56), however, they were also more inclined to use sunblock with a lower SPF (OR for a higher SPF 0.19; 95% CI 0.17, 0.22) and to apply it more often (OR for shorter application time 1.41, 95% CI 1.23, 1.61).

As for the reasons given for liking to tan, sex was strongly associated with every reason except for looking more attractive to others and thinking pale skin is unattractive. Higher parental social class and maternal education were inversely associated with thinking that pale skin is unattractive, but positively associated with tanning to look more attractive to others. Skin colour and tanning ability showed evidence of a strong association with thinking pale skin is unattractive, while individuals who experienced more sunburns in the past two years reported liking to tan because they look better in photos, look more attractive to others and also think pale skin is unattractive. Lastly, believing that tanning helps prevent sunburn was positively associated with all motives for tanning. (Supplementary Table 7).

#### 2. Outdoor tanning

Predictors of outdoor tanning are presented in Supplementary Table 8. Males were considerably more likely than females to tan outdoors (OR 6.11; 95% CI 3.29, 11.32). A positive association was also evident with parental higher SEP and maternal higher education. Skin colour, hair colour, tanning ability, having freckles, experiencing a painful sunburn in the past 2 years and believing that indoor tanning helps prevent sunburn were all factors associated with outdoor tanning, usually in the direction of individuals with darker phenotypes practicing this activity more often.

#### 3. Indoor tanning using UV equipment

Liking to tan was a strong predictor for ever using UV indoor tanning (OR 7.65; 95% CI 6.26, 9.34). Usually tanning indoors with a sunbed, sun lamp or tanning booth was strongly associated with female sex, but pigmentation and sun exposure traits showed weaker effects (Table 3). Additional associations were uncovered with parents’ SEP and maternal education such that individuals whose parents had a non-manual occupation and more educated mothers were less likely to engage in UV indoor tanning. Participants who believe that indoor tanning helps prevent sunburn were at a substantially increased risk of undertaking UV indoor tanning.

**Table 3.** Association of socio-demographic, pigmentation-related and sun exposure factors with usually tanning indoors using a sun bed/sun lamp/tanning booth amongst ALSPAC young people who like to tan.

| **exposure** | **OR<sup>a</sup>** | **95% CI** | **p-value** | **N** |
| --- | --- | --- | --- | --- |
| **sex** (0=female/1=male) | 0.47 | (0.33,0.66) | 1.10x10<sup>-5</sup> | 1930 |
| **age** (years) | 0.94 | (0.76,1.17) | 0.598 | 1930 |
| **mother's social class** (0=manual/1=non-manual) | 0.55 | (0.39,0.78) | 0.001 |  |
| **father's social class** (0=manual/1=non-manual) | 0.52 | (0.40,0.67) | 8.32x10<sup>-7</sup> | 1704 |
| **mother's education** |  |  | 4.31x10<sup>-6</sup> | 1814 |
| < O level | reference |  |  |  |
| O level | 0.98 | (0.70,1.36) | 0.885 |  |
| > O level | 0.50 | (0.36,0.70) | 6.70x10<sup>-5</sup> |  |
| **skin colour** |  |  | 0.002 | 1929 |
| very fair | reference |  |  |  |
| fair | 1.92 | (1.19,3.11) | 0.007 |  |
| olive | 2.60 | (1.57,4.30) | 2.00x10<sup>-4</sup> |  |
| brown | 2.10 | (0.95,4.64) | 0.066 |  |
| **eye colour** |  |  | 0.451 | 1926 |
| blue | reference |  |  |  |
| green | 1.07 | (0.78,1.48) | 0.673 |  |
| grey | 1.64 | (0.85,3.16) | 0.138 |  |
| brown | 1.09 | (0.81,1.47) | 0.566 |  |
| other | 0.72 | (0.37,1.40) | 0.336 |  |
| **natural hair colour at 18 years old** |  |  | 0.476 | 1927 |
| red | reference |  |  |  |
| blonde | 1.86 | (0.64,5.39) | 0.256 |  |
| light brown | 2.25 | (0.79,6.41) | 0.128 |  |
| dark brown | 2.25 | (0.79,6.41) | 0.129 |  |
| black | 1.20 | (0.12,11.75) | 0.877 |  |
| other | 1.31 | (0.27,6.37) | 0.741 |  |
| **YP has freckles** |  |  | 0.091 | 1926 |
| no | reference |  |  |  |
| yes, a few | 1.30 | (1.00,1.69) | 0.054 |  |
| yes, many | 0.95 | (0.62,1.43) | 0.793 |  |
| **tanning ability** |  |  | 0.001 | 1923 |
| always burns never tans | reference |  |  |  |
| burns easily rarely tans | 0.82 | (0.27,2.48) | 0.729 |  |
| doesn't change | 1.02 | (0.29,3.58) | 0.978 |  |
| tans easily rarely burns | 1.44 | (0.49,4.22) | 0.506 |  |
| always tans never burns | 2.47 | (0.76,7.97) | 0.132 |  |
| can't say, skin always protected | 0.69 | (0.16,3.03) | 0.619 |  |
| **sun burning in the past 2 years** |  |  | 0.118 | 1868 |
| never | reference |  |  |  |
| once | 0.66 | (0.46,0.94) | 0.021 |  |
| twice | 0.66 | (0.47,0.93) | 0.019 |  |
| 3 times | 0.66 | (0.41,1.04) | 0.074 |  |
| 4 times | 0.82 | (0.48,1.38) | 0.448 |  |
| 5 times or more | 0.99 | (0.55,1.78) | 0.973 |  |
| **sun protection (0=no/1=yes)** |  |  |  |  |
| never uses protection | 1.56 | (1.01,2.40) | 0.046 | 1928 |
| wears hat | 0.84 | (0.62,1.13) | 0.255 | 1928 |
| wears clothing | 0.56 | (0.41,0.78) | 0.001 | 1928 |
| wears sunblock | 0.67 | (0.42,1.05) | 0.081 | 1928 |
| avoids the sun | 0.35 | (0.19,0.65) | 0.001 | 1928 |
| **sunblock factor** |  |  | 4.20x10<sup>-12</sup> | 1798 |
| lower than 15 | reference |  |  |  |
| 15-24 | 0.43 | (0.29,0.64) | 3.08x10<sup>-5</sup> |  |
| 25-49 | 0.23 | (0.15,0.35) | 6.11x10<sup>-12</sup> |  |
| 50 or higher | 0.17 | (0.08,0.35) | 1.06x10<sup>-6</sup> |  |
| **skin cancer** (0=no/1=yes) |  |  |  |  |
| YP has family member diagnosed with skin cancer | 0.92 | (0.63,1.32) | 0.640 | 1925 |
| **beliefs about tanning** |  |  |  |  |
| YP believes indoor tanning helps prevent sunburn |  |  | 2.25x10<sup>-39</sup> | 1926 |
| no | reference |  |  |  |
| yes | 8.45 | (6.16,11.59) | < 10<sup>-26</sup> |  |
| don't know | 1.47 | (1.04,2.07) | 0.029 |  |
| YP thinks indoor tanning using sun bed/sun lamp can cause skin cancer |  |  | 0.432 | 1924 |
| no | reference |  |  |  |
| yes | 0.65 | (0.33,1.26) | 0.198 |  |
| don't know | 0.68 | (0.32,1.46) | 0.324 |  |
| **body image** |  |  |  |  |
| YP is trying to change skin colour (0=no/1=yes) | 2.07 | (1.42,3.03) | 1.74x10<sup>-4</sup> | 1537 |
| YP is trying to change hair colour (0=no/1=yes) | 2.05 | (1.32,3.20) | 0.001 | 1537 |
| YP has been hurt because of their skin colour (0=no/1=yes) | 0.65 | (0.08,5.20) | 0.683 | 1604 |
| YP has been called names because of their skin colour (0=no/1=yes) | 0.84 | (0.29,2.46) | 0.754 | 1608 |
| YP has seen someone bullied because of their skin colour (0=no/1=yes) | 1.34 | (1.00,1.79) | 0.049 | 1599 |
| YP's teachers treat everybody the same, regardless of skin colour (0=agree/1=disagree) | 1.36 | (0.67,2.76) | 0.390 | 1478 |

<sup>a</sup>Adjusted for age and sex.

We also examined determinants of recent indoor tanning using a sunbed (i.e. in the past 12 months) (Supplementary Table 9). Notable positive associations were found with lower father’s SEP, darker skin colour, good tanning ability, use of sunblock with low SPF, and believing that indoor tanning helps prevent sunburn. Furthermore, we found evidence that subjects who thought that indoor tanning can cause skin cancer were less likely to have used a sunbed recently (Supplementary Table 9).

#### 4. Indoor tanning using non-UV methods

Fifty percent of individuals who declared usually UV tanning indoors used sunless tanning products as well. Indoor tanning with either a spray tan or self-tanning lotions was associated with sex, father’s social class and mother’s education in the same direction as usually indoor tanning using a sunbed. But skin colour, tanning ability and having freckles exhibited associations in the opposite direction, i.e. individuals with very fair skin, who always burn, and have freckles, were more likely to go for a spray tan or use self-tanning lotions than individuals with darker phenotypes, who were keener to use a sunbed. Supplementary Table 10.

#### 5. Body image and tanning

Although YP who reported trying to change their skin and hair colour at ∼14 years old did not manifest a preference for tanning (Table 2), those who liked to tan, were more inclined to indoor tan using a UVR device (Table 3) or a non-UV method (Supplementary Table 10) later in life. Similar associations were found with having ever used a sunbed (OR 1.90, 95% CI 1.46, 2.98 for changing skin colour; OR 1.59, 95% CI 1.17, 2.16 for changing hair colour), but not with having used a sunbed in the past 12 months (Supplementary Table 9). We tested whether skin reflectance, tanning ability and hair colour measured or informed before 14 years of age could explain the willingness to change skin or hair colour in adolescence and found no robust associations. Notably, the desire to change appearance was strongly associated with being a female (Supplementary Table 11).

There were positive associations with several reasons for liking to tan, more so among participants who had tried to change their skin colour (Supplementary Table 7).

YP who disagreed with the statement about teachers treating everybody the same way, regardless of skin colour, favoured undergoing non-UV tanning (OR 2.30, 95% CI 1.21, 4.37) (Supplementary Table 10). They were also more likely to select ‘pale skin is unattractive’ as a reason for tanning (OR 2.04, 95% CI 1.15,3.63) (Supplementary Table 7).

Finally, children who reported being hurt or having been called names because their skin colour was different than that of their peers, and children who had seen someone being bullied because of their skin colour, exhibited lower odds of liking to tan (Table 2). The former were also less likely to have ever used a sunbed, although the evidence for association was weak (OR 0.55, 95% 0.21, 1.47 and OR 0.53, 95% 0.28, 1.01, respectively).

### Genetically-informed analysis

We checked that the probabilities estimated with HIrisPlex-S were in fact correlated with the pigmentation traits they were expected to predict. In addition, their association with sun exposure-related variables, such as tanning ability, sun burning or sun protection was tested. Strong associations were found throughout (see Supplementary Tables 12 and 13).

HIrisPlex-S estimates were not correlated with most principal components, although PC6 showed a weak correlation with eye and hair colour prediction probabilities, and so did PC10 with the prediction probability of skin colour (Supplementary Table 14).

Despite blond and brown self-reported hair colour being unevenly distributed in ALSPAC males and females, in agreement with earlier studies of British and European populations^33, 34^ there were no differences by sex in the genetic probabilities for each trait (blond hair: OR 0.93, 95% CI 0.77, 1.13; brown hair: OR 0.93, 95% CI 0.74, 1.18; black hair: OR 1.01, 95% CI 0.61,1.69; reference: female sex).

#### 1. Genetically-determined pigmentation traits vs hair and eye colour text answers

We tested whether the hair and eye colour categories defined by the text analysis had any genetic basis. We identified strong positive associations of the text answer categories of red and ginger hair with the probability of having red hair. The blue eye text category was positively associated with the probability of having blue-coloured eyes, and so was the green eye category, although with a weaker effect. Interestingly, the category hazel eyes was strongly and positively associated with the probability of having eyes of intermediate colour and the probability of having brown eyes (though not as strongly), and inversely with the probability of having blue eyes. There was not enough evidence of an association of genetically-determined hair and eye colour with any of the other text categories (Supplementary Table 15).

#### 2. Genetically-determined pigmentation traits vs tanning preferences

Genetically-determined pigmentation traits were robustly associated with liking to tan in the same way uncovered in the observational study. Brown eyes, black hair, intermediate skin and dark skin showed a positive association with liking to tan, whereas blue eyes, red hair, pale skin and very pale skin had the opposite effect (Table 4). There were also positive associations of black hair and intermediate skin, and inverse associations of red hair and very pale skin, with outdoor tanning (Table 4), but no clear evidence of an effect of pigmentation on indoor tanning with a UVR device (Table 5). Adjusting the regression models for 5 PCs only did not change the latter results appreciably. Conversely, genetically-determined very pale skin and, to a lesser extent, red hair, increased the odds of using sunless tanning products (Supplementary Table 16).

**Table 4.**
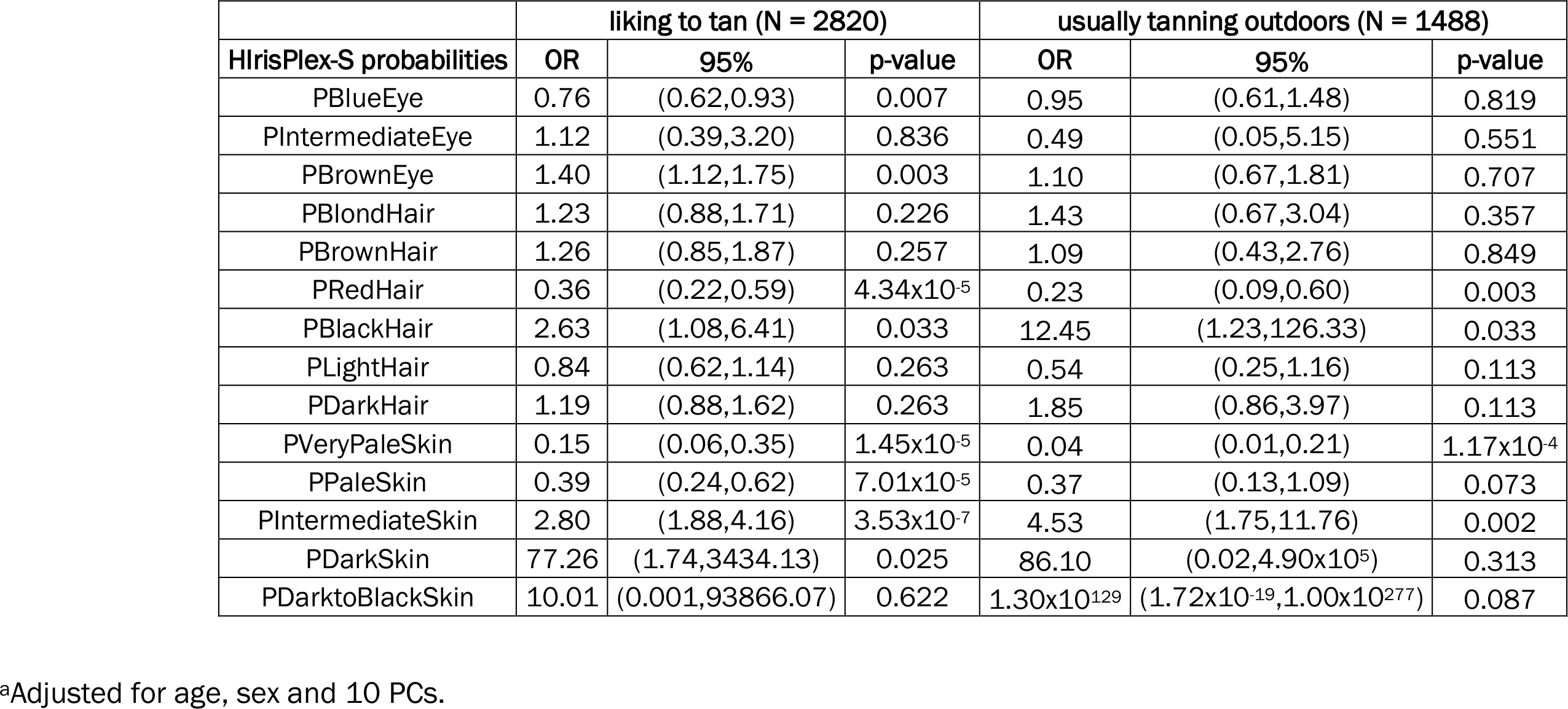
Association of liking to tan and usually tanning outdoors with HIrisPlex-S prediction probabilities of pigmentation traits in ALSPAC young people.

**Table 5.**
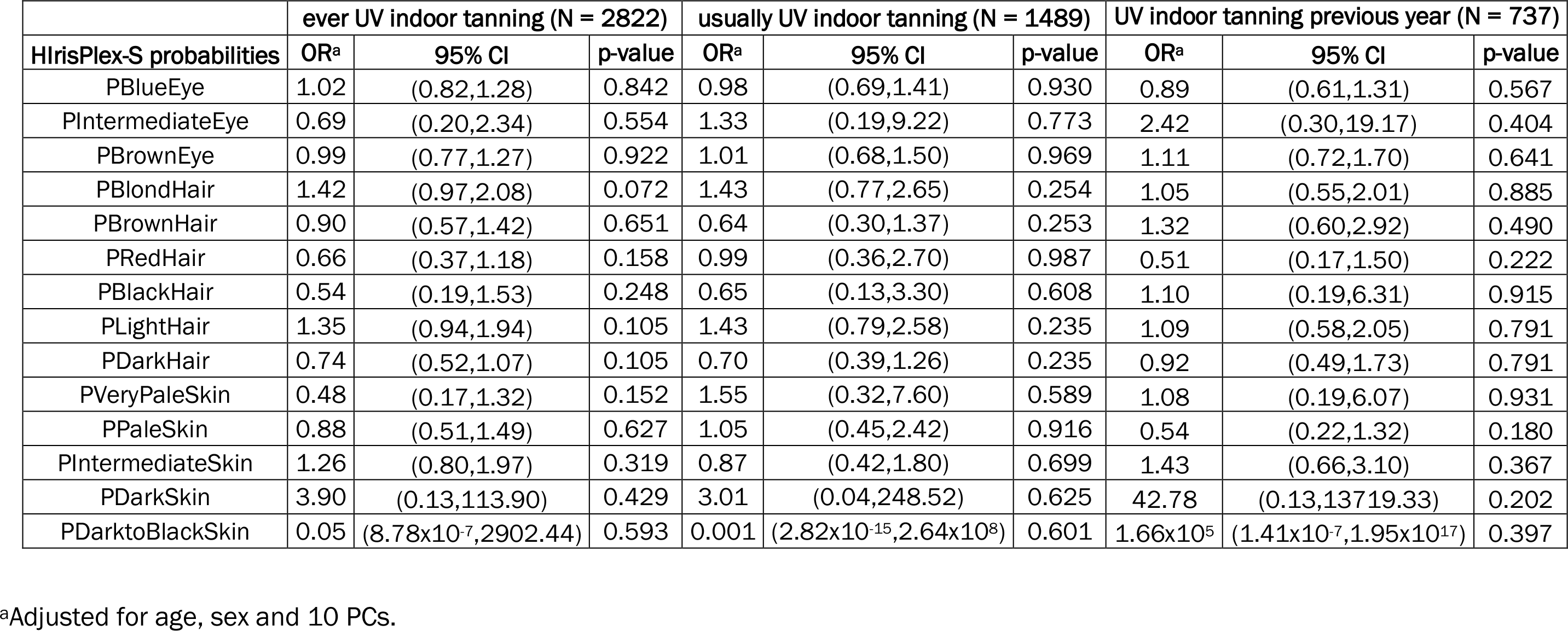
Association of having ever used UV indoor tanning equipment, usually undergoing UV indoor tanning and UV indoor tanning in the last 12 months with HIrisPlex-S prediction probabilities of pigmentation traits in ALSPAC young people.

|  | ever UV indoor tanning (N = 2822) |  |  | usually UV indoor tanning (N = 1489) |  |  | UV indoor tanning previous year (N = 737) |  |  |
| --- | --- | --- | --- | --- | --- | --- | --- | --- | --- |
| HirisPlex-S probabilities | OR<sup>a</sup> | 95% CI | p-value | OR<sup>a</sup> | 95% CI | p-value | OR<sup>a</sup> | 95% CI | p-value |
| PBlueEye | 1.02 | (0.82,1.28) | 0.842 | 0.98 | (0.69,1.41) | 0.930 | 0.89 | (0.61,1.31) | 0.567 |
| PIntermediateEye | 0.69 | (0.20,2.34) | 0.554 | 1.33 | (0.19,9.22) | 0.773 | 2.42 | (0.30,19.17) | 0.404 |
| PBrownEye | 0.99 | (0.77,1.27) | 0.922 | 1.01 | (0.68,1.50) | 0.969 | 1.11 | (0.72,1.70) | 0.641 |
| PBlondHair | 1.42 | (0.97,2.08) | 0.072 | 1.43 | (0.77,2.65) | 0.254 | 1.05 | (0.55,2.01) | 0.885 |
| PBrownHair | 0.90 | (0.57,1.42) | 0.651 | 0.64 | (0.30,1.37) | 0.253 | 1.32 | (0.60,2.92) | 0.490 |
| PRedHair | 0.66 | (0.37,1.18) | 0.158 | 0.99 | (0.36,2.70) | 0.987 | 0.51 | (0.17,1.50) | 0.222 |
| PBlackHair | 0.54 | (0.19,1.53) | 0.248 | 0.65 | (0.13,3.30) | 0.608 | 1.10 | (0.19,6.31) | 0.915 |
| PLightHair | 1.35 | (0.94,1.94) | 0.105 | 1.43 | (0.79,2.58) | 0.235 | 1.09 | (0.58,2.05) | 0.791 |
| PDarkHair | 0.74 | (0.52,1.07) | 0.105 | 0.70 | (0.39,1.26) | 0.235 | 0.92 | (0.49,1.73) | 0.791 |
| PVeryPaleSkin | 0.48 | (0.17,1.32) | 0.152 | 1.55 | (0.32,7.60) | 0.589 | 1.08 | (0.19,6.07) | 0.931 |
| PPaleSkin | 0.88 | (0.51,1.49) | 0.627 | 1.05 | (0.45,2.42) | 0.916 | 0.54 | (0.22,1.32) | 0.180 |
| PIntermediateSkin | 1.26 | (0.80,1.97) | 0.319 | 0.87 | (0.42,1.80) | 0.699 | 1.43 | (0.66,3.10) | 0.367 |
| PDarkSkin | 3.90 | (0.13,113.90) | 0.429 | 3.01 | (0.04,248.52) | 0.625 | 42.78 | (0.13,13719.33) | 0.202 |
| PDarktoBlackSkin | 0.05 | (8.78x10<sup>-7</sup>,2902.44) | 0.593 | 0.001 | (2.82x10<sup>-15</sup>,2.64x10<sup>8</sup>) | 0.601 | 1.66x10<sup>5</sup> | (1.41x10<sup>-7</sup>,1.95x10<sup>17</sup>) | 0.397 |

<sup>a</sup>Adjusted for age, sex and 10 PCs.

Among the reasons for liking to tan thinking pale skin is unattractive was associated with genetically determined pale skin (positively) and dark skin (inversely), as expected (Supplementary Table 17). HIrisPlex-S pigmentation probabilities were not associated with any other reasons for tanning.

YP with genetically predicted blue eyes and pale skin were less likely to expose themselves to the sun unprotected, while those with predicted brown eyes, intermediate skin and dark skin were more prone to do so. In general, higher levels of protection, be it wearing a hat, clothing, sunblock or directly avoiding the sun, were more widespread among individuals with genetically determined blue eyes, red hair, pale skin and very pale skin, and less common among YP with genetically determined brown eyes, black hair, intermediate skin and dark skin. A similar trend was observed with respect to using sunblock with higher SPF. Remarkably, participants with predicted blond hair were less inclined to wear clothing or avoid the sun as a protective measure (Supplementary Table 18).

Trying to change skin colour to match media images was more likely in individuals with genetically determined brown eyes and black/dark hair, and less probable in subjects with genetically determined blond/light hair. Children with likely brown eyes, black hair and dark skin reported having been called names because their skin colour was different more frequently, as opposed to children with likely pale skin. These associations were not present among children who declared having been hurt because of their different skin colour (Supplementary Table 19).

## Discussion

We performed an epidemiological study informed by survey and genetic data to ascertain socio-demographic and pigmentation-related determinants of outdoor and indoor tanning behaviour in young adults of European ancestry (ALSPAC participants of ∼25 years of age). In particular, we were interested in their answers to the question “do you like to tan?” because it identifies individuals who have the motivation or propensity to deliberately expose themselves to UVR. Although all respondents were of approximately the same age, there was diversity with respect to their SEP growing up, allowing us to investigate tanning preferences across a wider socioeconomic spectrum than studies that were limited to university students.

We found that women were considerably more likely than men to say they liked to tan. In agreement with earlier studies, they were more inclined to engage in indoor tanning either using a sunbed^7, 35^, or a non-UV method, such as a spray tan or self-tanning lotions^36^. On the other hand, men reported tanning outdoors more frequently than women, which is also in line with the literature^22^.

Overall, having a darker pigmentation phenotype increased the odds that participants were fond of tanning. But YP who had experienced several sunburns in the past two years reported enjoying tanning more than those who had not suffered them, which, although it may appear contradictory to what their tanning ability would suggest, implies that their desire for a tan overrides the harm that it may cause. Indeed, the main reasons these individuals provided for liking to tan are looking better in photos and looking more attractive to others.

The analysis of pigmentary traits and indoor tanning by Li et al. on the Nurses’ Health Study (NHS)^6^, a large prospective cohort, showed that women with light hair colour (blond/red) were keener to use a sunbed than women with brown/black hair colour. In our study participants with blond and red hair differed in their tanning inclinations, with blond hair YP exhibiting a more favourable approach to tanning and reporting use of a sunbed in greater numbers than YP with red hair. The NHS study also found that women who experienced more severe sunburns as teenagers tanned indoors more frequently, whilst women with lower tanning ability in childhood preferred to avoid this activity^6^.

In terms of the reasons given for liking to tan, the most frequent were related to confidence, happiness, and attractiveness. Improving attractiveness has been a fairly common motive for tanning across studies^37–39^. Additionally, more than 20% of respondents who liked to tan mentioned pale skin being unattractive as a reason for tanning, and therefore, individuals with darker skin and a better tanning ability were substantially less inclined to support this answer. In contrast, participants who suffered episodes of sun burning, do not use sun protection, and who thought their teachers did not treat everybody the same regardless of skin colour, were more likely to refer to the unattractiveness of pale skin as a motive for tanning.

Individuals who tried to modify their hair or skin colour to match media messages were prone to UV and non-UV indoor tan, even though they did not exhibit a greater preference for tanning than YP who did not attempt those changes. Trying to change skin colour in early adolescence was positively associated with all reasons for liking to tan, suggesting that feeling better about oneself and looking more attractive to others are all encompassing aims of tanning in this group of people. Our findings could potentially be understood under socio-cultural body image theories where tanning may be driven by the positive aspect of becoming more attractive but also by the negative effects of not fitting a tan beauty ideal, leading to body dissatisfaction^21^. As a matter of fact, in a study run in the context of a potential advertising campaign, the author found that female models with a light brown skin tone were perceived by European Americans and African Americans as being more credible and physically attractive than those with pale or dark skin tones^40^. Another study, carried out in college students in the UK, found that hair colour was an important variable in interpersonal judgements of attractiveness, more so than skin tone and hair length^41^.

With respect to sun protection, we identified three distinctive groups of participants: participants who were very conscientious about it (i.e. avoiding the sun and/or wearing clothing or a hat) and frequently reported not liking to tan; participants who liked to tan and their strategy of protection consisted in applying sunblock repeatedly but with a lower SPF; and participants who liked to tan and never protected themselves in the sun (Table 2). The identification of these differential behaviours is valuable to implement preventive skin cancer campaigns, specially targeted to the latter two groups.

A diagnosis of skin cancer, whether personal or of a family member, did not seem to influence YP’s tanning predilection, although sample sizes were small for this analysis. In addition, awareness of UV indoor tanning causing skin cancer did not show an effect on liking to tan, but reduced the odds that these individuals have used a sunbed in the past 12 months. Remarkably, ∼85% of YP who answered the Life@25+ questionnaire were knowledgeable about the skin cancer risks of indoor tanning, thus, as others have also observed^22, 23^, individuals who still engage in this behaviour do so because they assess its benefits as being worth the risks. This contradiction may be explained by what is known as “optimistic bias”, where a person believes that the harmful consequences of a particular action are less likely to happen to them than to others^42^. It also suggests that interventions aimed at increasing knowledge about the dangers of excessive UVR exposure are unlikely to induce a behavioural change among individuals with powerful motivations for tanning. On the other hand, a non-trivial fraction of participants (12%) responded that they did not know whether the skin cancer assertion was true, with 3% denying the existence of risk.

Particular consideration should be given to individuals who believe that indoor tanning helps prevent sunburn, despite representing just ∼8% of all respondents, as they are at an appreciably increased risk of engaging in indoor tanning using a sunbed, and having used indoor tanning equipment in the past year. This finding agrees with a French study that described higher odds of indoor tanning for these subjects (OR 4.3, 95% CI 2.7, 6.7)^19^, and similarly, with the 2014 and 2019 Canadian Community Health Surveys^43^,^44^.

Usually tanning indoors using a sunbed as well as having used indoor tanning equipment in the past 12 months was less frequent among participants with higher SEP, association that has been reported previously in the same^7, 20^ and opposite directions^19, 43^. Darker skinned individuals, with good tanning ability, and who were unlikely to use sun protection were found to engage more often in this activity and to have done so in the past year. Similar results were uncovered by Haluza et al. in Austria^45^ and Grange et al.^46^ in France.

Since there was substantial overlap between participants who opted for different indoor tanning methods, as has been shown for other populations^36, 45, 47^, the promotion of the “healthier” option of a spray tan or self-tanning lotions for UV indoor tanners constitutes a potentially useful intervention, especially in view of persistent tanning as an addictive behaviour^48–50^. This possibility has already been brought up by the American Academy of Dermatologists (https://www.aad.org/public/diseases/skin-cancer/surprising-facts-about-indoor-tanning) and Cancer Research UK (https://www.cancerresearchuk.org/about-cancer/causes-of-cancer/sun-uv-and-cancer/fake-tan-and-melanotan-injections). Nevertheless, it appears that individuals with more sensitive skins, who are at an increased risk for skin cancer, are already choosing these alternatives. Equally, there may be some health adverse consequences associated with “fake tanning” that need to be further explored^36^. Diehl et al. found that subjects who employed sunless tanning products had less knowledge about the health risks posed by indoor tanning^36^. In our study, participants who believed that UV indoor tanning helps prevent sunburn showed higher odds of tanning using non-UV methods (OR 1.84; 95% CI 1.35, 2.52).

In our genetically-informed analysis, we identified putative causal associations between pigmentation traits and tanning behaviour. Genotypes are set at conception and are generally not affected by various biases that plague observational studies, including confounding, response and recall bias, and reverse causality. Self-reported burning tendencies and tanning ability may be inconsistent as well^52, 53^.

We found that liking to tan and outdoor tanning were clearly affected by skin, hair and eye pigmentation, but that was not the case with indoor tanning using a UVR device. It is possible that UV indoor tanning is controlled by a number of different factors and that pigmentation is not comparatively as relevant as other determinants. For instance, it has been reported that indoor tanning is associated with mental health problems^54^, physical activity and dietary practices^55^, and other risky behaviours^56, 57^. Moreover, a recent GWAS meta-analysis on sun-seeking behaviour^58^, adjusted for tanning ability and SEP, identified 5 loci related to behavioural traits, including addiction. In our study, additional factors included beliefs about tanning, which were unrelated to genetically-determined pigmentation traits (Supplementary Table 20), and parental education and SEP. Identifying the causes of indoor tanning behaviour warrants further investigation to inform policy and practice.

Lastly, it would be valuable to assess the consequences that tanning preferences may have on vitamin D levels. For example, 3.8% of individuals who responded the tanning ability question mentioned that their skin is always covered (Table 1). Although ALSPAC has measured circulating vitamin D when YP were 7, 9 and 11 years old, and we found an association of pigmentation genetic scores with these vitamin D levels in an earlier study^30^, the impact of adult decisions and motivations towards sun exposure on serum vitamin D concentrations remains to be established.

## Limitations

Our observational analysis may be affected by confounding and reverse causation, especially since a number of exposures and outcomes were obtained cross-sectionally, and there may be unmeasured confounders at play. Additionally, the questionnaire we applied did not include questions about peer-pressure and sociocultural influences that may explain, at least in part, tanning choices made by the participants in the study^23^.

Our genetically-informed analysis was conducted using the HIrisPlex-S system, developed and validated in European populations, therefore applicability of our findings to non-European populations might not be appropriate. Recently, genetic variants associated with pigmentation traits have been discovered in populations of other ancestries^59^. Although a similar predictive algorithm is not available for them yet, HIrisPlex-S (or its related versions: HIrisPlex and IrisPlex) has been tested in some Latin American and Asian populations, showing an acceptable performance for eye and hair colour predictions, better than for skin pigmentation, but exhibiting limitations in the prediction of intermediate phenotypes, which are more frequent in these groups^60–63, 64, 65^. Future research that addresses these shortcomings will not only advance the field of forensics but also will facilitate genetically-informed studies such as this one in admixed populations. Lastly, since we carried out our study in British individuals of European descent, HIrisPlex-S predictions of dark skin and dark to black skin may be unreliable, as point estimates showed very wide 95% CIs, therefore, they should be considered with caution.

## Conclusions

We uncovered that about half of ALSPAC YP who answered the Life@25+ questionnaire liked to tan and readily engaged in outdoor tanning. This preference was determined in large part by pigmentation traits. A quarter of respondents had ever used an indoor tanning UVR device, but those who usually underwent UV tanning were ∼8% of the study population.

Women liked to tan more than men and were more inclined to undertake UV indoor tanning, whereas men experienced sun exposure mostly outdoors. Attractiveness, happiness and confidence were the main reasons given for liking to tan. Individuals who believed that indoor tanning helps prevent sunburn were highly likely to practice all types of tanning and to do so with inadequate protection.

Future skin cancer preventative interventions among British youth, and possibly other populations of European ancestry, could focus on a message of satisfaction with their own skin (in fact, embracing their own appearance was one of the reasons given by former tanners to quit indoor tanning^66^), emphasize opposition to discrimination and stigmatization based on skin colour, reinforce the notion that sun burning, a skin cancer risk factor, is not preventable by tanning, and remind darker-skinned individuals of the risks of too much UVR exposure. Tailoring interventions by sex could also help. At the same time, it should be stressed that sun exposure can be responsibly enjoyed as there are many health benefits to be obtained from experiencing sunlight.

## Supplementary Tables legends

Supplementary Table 1. Best AUC at a population level and mean (standard deviation) loss of AUC for each predicted trait in ALSPAC young people (N = 8564).

Supplementary Table 2. Text answers of ALSPAC young people who responded “other reason” to the question “Why do you like to tan?”. Percentages over the total number of answers are shown.

Supplementary Table 3. Text answers of ALSPAC young people (YP) who reported having “other” hair colour. Percentages over the total number of answers are shown.

Supplementary Table 4. Text answers of ALSPAC young people who reported having “other” eye colour. Percentages over the total number of answers are shown.

Supplementary Table 5. Correlation between pigmentation, sun exposure and sun protection traits in childhood and the same or similar traits reported in the questionnaire Life@25+ for ALSPAC young people.

Supplementary Table 6. Text answers of ALSPAC young people who reported taking “other” measures for sun protection. Percentages over the total number of answers are shown.

Supplementary Table 7. Factors associated with reasons for liking to tan. Only those with the strongest evidence for association are shown.

Supplementary Table 8. Association of socio-demographic, pigmentation-related and sun exposure factors with outdoor tanning in ALSPAC young people who like to tan.

Supplementary Table 9. Association of socio-demographic, pigmentation-related and sun exposure factors with indoor tanning in the past 12 months amongst ALSPAC young people who manifested ever engaging in indoor tanning using a sun bed/sun lamp/tanning booth.

Supplementary Table 10. Association of socio-demographic, pigmentation-related and sun exposure factors with indoor tanning using spray tan or self-tanning lotions amongst ALSPAC young people who like to tan.

Supplementary Table 11. Association of skin reflectance, tanning ability and hair colour measured or informed before 14 years of age with trying to change skin and hair colour at age 14.

Supplementary Table 12. Pigmentation traits vs HIrisPlex-S prediction probabilities in ALSPAC young people (YP). Skin reflectance was obtained from clinic data collected when YP were 49 months old. All other pigmentation traits were obtained from the Life@25+ questionnaire.

Supplementary Table 13. Sun exposure traits vs HIrisPlex-S prediction probabilities in ALSPAC young people. All sun exposure traits were obtained from the Life@25+ questionnaire.

Supplementary Table 14. Correlation between HIrisPlex-S prediction probabilities and the top 10 genetic principal components (PCs) in ALSPAC young people.

Supplementary Table 15. Association of text answer categories of hair and eye colour with HIrisPlex-S prediction probabilities in ALSPAC young people.

Supplementary Table 16. Association of indoor tanning using sprays or self-tanning lotions with HIrisPlex-S prediction probabilities of pigmentation traits in ALSPAC young people.

Supplementary Table 17. Association of reasons for liking to tan with HIrisPlex-S prediction probabilities of pigmentation traits in ALSPAC young people.

Supplementary Table 18. Association of sun protection strategies with HIrisPlex-S prediction probabilities of pigmentation traits in ALSPAC young people.

Supplementary Table 19. Association of body image-related variables with HIrisPlex-S prediction probabilities of pigmentation traits in ALSPAC young people.

Supplementary Table 20. Association of beliefs about tanning with HIrisPlex-S prediction probabilities of pigmentation traits in ALSPAC young people.

## Supplementary Figure legends

Supplementary Figure 1. Connections between text answer categories of reasons for liking to tan reported by ALSPAC young people in the Tanning and Sun Exposure section of the Life@25+ questionnaire. Statistically significant coincidences are represented by dotted lines (p < 0.05) or a continuous line (p < 0.001).

Supplementary Figure 2. Skin and hair colour distribution among ALSPAC young people who completed the Tanning and Sun Exposure section of the Life@25+ questionnaire.

Supplementary Figure 3. Skin and eye colour distribution among ALSPAC young people who completed the Tanning and Sun Exposure section of the Life@25+ questionnaire.

Supplementary Figure 4. Hair and eye colour distribution among ALSPAC young people who completed the Tanning and Sun Exposure section of the Life@25+ questionnaire.

Supplementary Figure 5. Hierarchical clustering visualization of sun protection strategies reported as text answers by ALSPAC young people in the Tanning and Sun Exposure section of the Life@25+ questionnaire.

## Data Availability

All data produced in the present study are available upon reasonable request to the authors.

## Acknowledgments

We are extremely grateful to all the families who took part in this study, the midwives for their help in recruiting them, and the whole ALSPAC team, which includes interviewers, computer and laboratory technicians, clerical workers, research scientists, volunteers, managers, receptionists and nurses.

## Funding

CB was supported by a Universidade de São Paulo/ Coordenação de Aperfeiçoamento de Pessoal de Nível Superior fellowship (88887.160006/2017-00), and a grant from the Pró-Reitoria de Pesquisa of the University of São Paulo (775/2020).

The UK Medical Research Council and Wellcome (Grant ref: 217065/Z/19/Z) and the University of Bristol provide core support for ALSPAC. GWAS data was generated by Sample Logistics and Genotyping Facilities at Wellcome Sanger Institute and LabCorp (Laboratory Corporation of America) using support from 23andMe. A comprehensive list of grant funding is available on the ALSPAC website (http://www.bristol.ac.uk/alspac/external/documents/grant-acknowledgements.pdf).

This publication is the work of the authors and Carolina Bonilla will serve as guarantor for the contents of this paper.

## Authors’ contributions

CB conceived the study, carried out the analysis and wrote the original draft. CM-L provided expertise in epidemiology and text analysis. Both authors critically revised the manuscript.

