## Supplementary Figures 1-5 for "The skin we live in: pigmentation traits and tanning behaviour in British young adults, an observational and genetically-informed study"

**Supplementary Figure 1.** Connections between text answer categories of reasons for liking to tan reported by ALSPAC young people in the Tanning and Sun Exposure section of the Life@25+ questionnaire. Statistically significant coincidences are represented by dotted lines ($p < 0.05$) or a continuous line ($p < 0.001$).

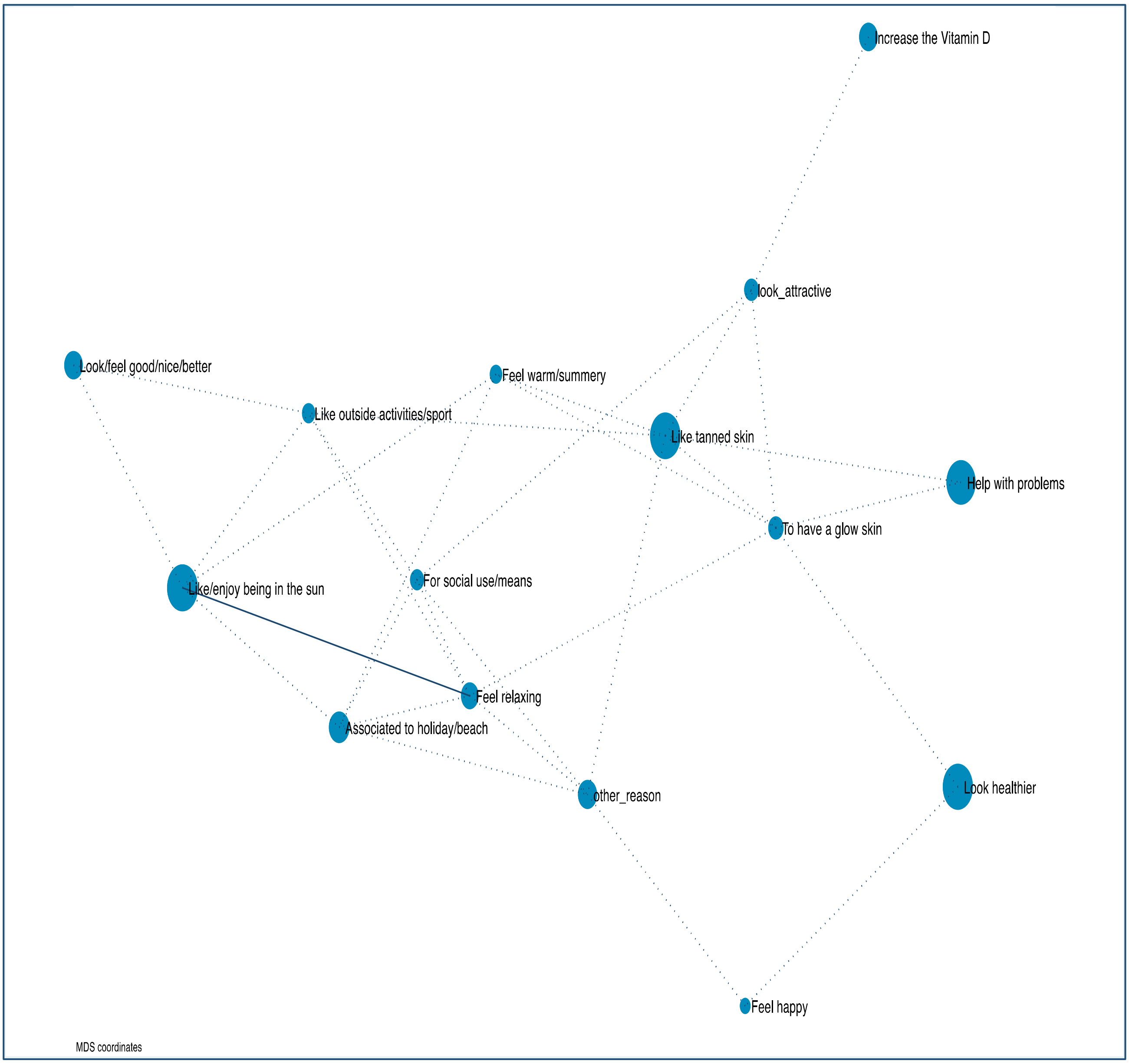

**Supplementary Figure 2.** Skin and hair colour distribution among ALSPAC young people who completed the Tanning and Sun Exposure section of the Life@25+ questionnaire.

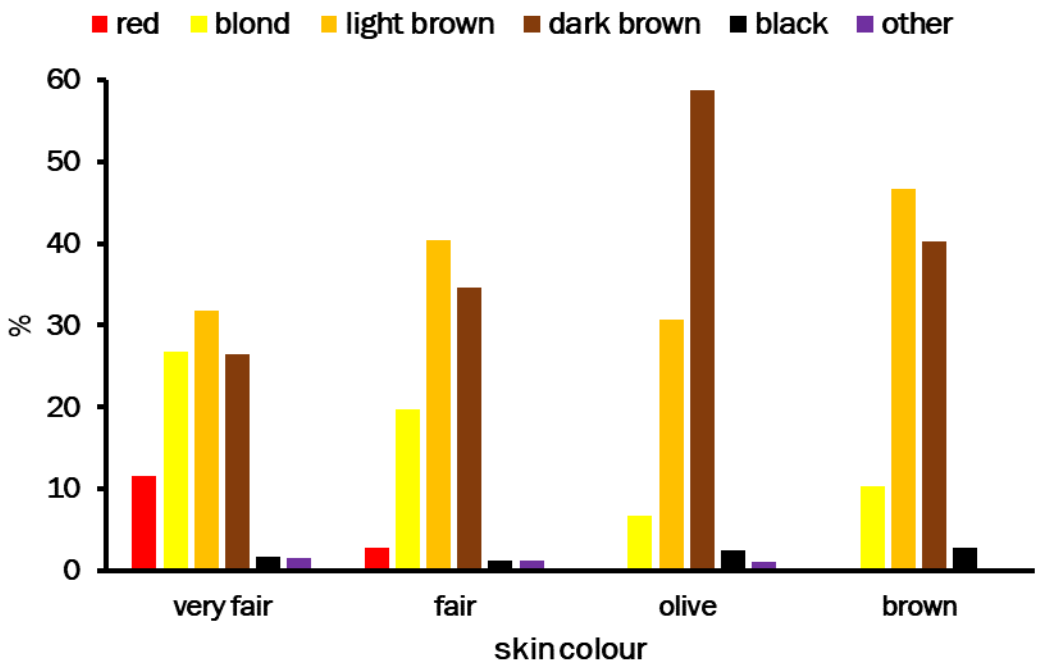

**Supplementary Figure 3.** Skin and eye colour distribution among ALSPAC young people who completed the Tanning and Sun Exposure section of the Life@25+ questionnaire.

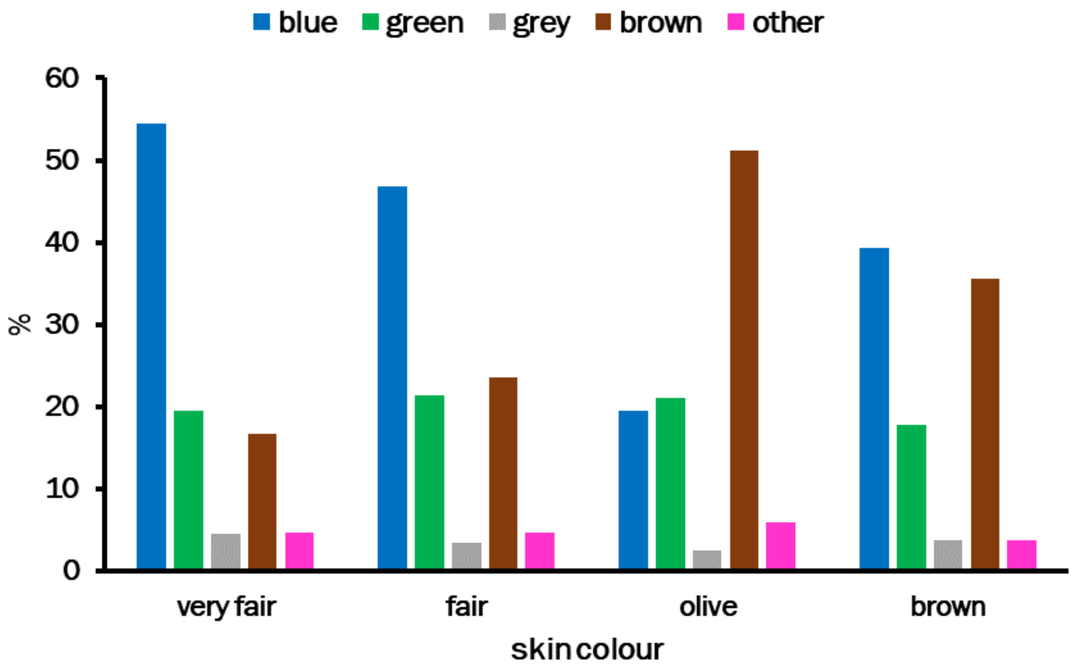

**Supplementary Figure 4.** Hair and eye colour distribution among ALSPAC young people who completed the Tanning and Sun Exposure section of the Life@25+ questionnaire.

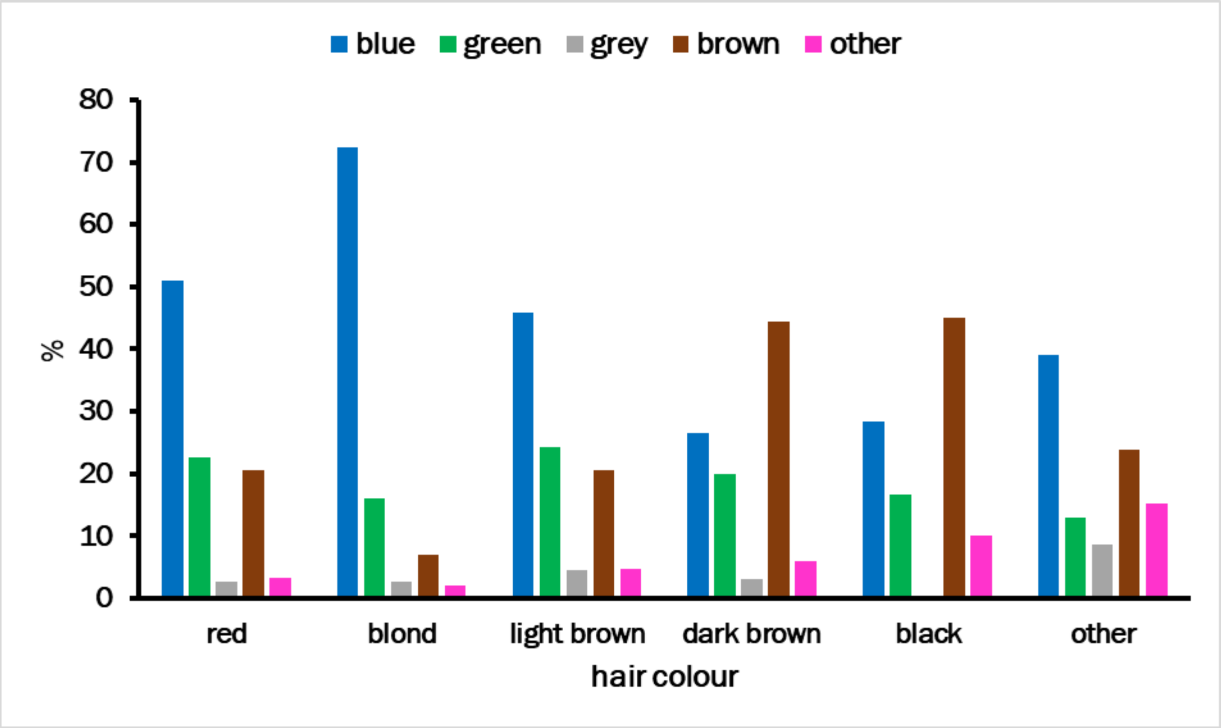

**Supplementary Figure 5.** Hierarchical clustering visualization of sun protection strategies reported as text answers by ALSPAC young people in the Tanning and Sun Exposure section of the Life@25+ questionnaire.

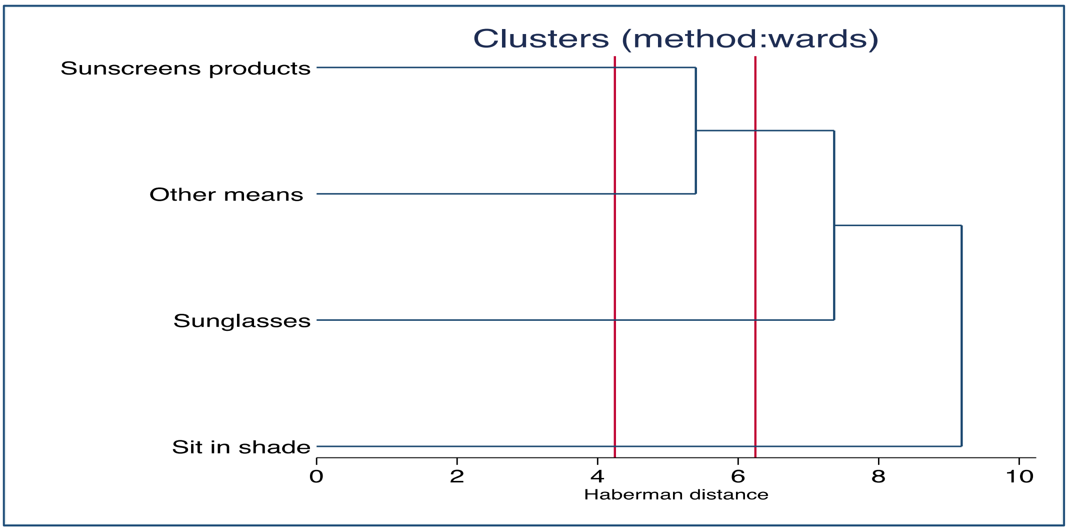
