## Supplementary Tables 1-4, 6-10, 16 for "The skin we live in: pigmentation traits and tanning behaviour in British young adults, an observational and genetically-informed study"

**Supplementary Table 1.** Best AUC at a population level and mean (standard deviation) loss of AUC for each predicted trait in ALSPAC young people (N = 8564).

| trait | full AUC | loss of AUC |
|---|---|---|
| blue eyes | 0.939 | 0.00042 (0.00098) |
| intermediate eyes | 0.736 | 0.00172 (0.00398) |
| brown eyes | 0.946 | 0.00071 (0.00167) |
| blond hair | 0.813 | 0.00403 (0.00445) |
| brown hair | 0.741 | 0.00252 (0.00390) |
| red hair | 0.929 | 0.00858 (0.01692) |
| black hair | 0.859 | 0.00196 (0.00341) |
| hair shade | 0.905 | 0.00160 (0.00297) |
| very pale skin | 0.830 | 0.00101 (0.00314) |
| pale skin | 0.763 | 0.00225 (0.00335) |
| intermediate skin | 0.783 | 0.00307 (0.00434) |
| dark skin | 0.981 | 0.00087 (0.00207) |
| dark to black skin | 0.993 | 0.00051 (0.00081) |

**Supplementary Table 2.** Text answers of ALSPAC young people who responded "other" to the question "Why do you like to tan?". Percentages over the total number of answers are shown.

| reasons | n = 331 answers | % |
|---|---|---|
| like or enjoy being under the sun | 49 | 14.8 |
| like tanned skin | 48 | 14.5 |
| look or be healthier | 47 | 14.2 |
| helps with skin problems such as acne, eczema, spots | 44 | 13.3 |
| associated to holidays or beach | 21 | 6.3 |
| increase vitamin D | 17 | 5.2 |
| look or feel good/nice | 17 | 5.2 |
| is relaxing/enjoyable | 15 | 4.5 |
| social reasons & professional reasons | 12 | 3.6 |
| have glowing skin | 11 | 3.3 |
| look more attractive | 10 | 3.0 |
| due to outside activities/sports | 8 | 2.4 |
| feel warm or summery | 7 | 2.1 |
| feel happy | 5 | 1.5 |
| other reasons (YP is naturally tanned, just happens, likes freckles, no special reason, does it sometimes, why not do that, photos/shoots, does not do it frequently) | 20 | 6.0 |

**Supplementary Table 3.** Text answers of ALSPAC young people who reported having "other" hair colour. Percentages over the total number of answers are shown.

| hair colour category | n = 79 answers | % |
|---|---|---|
| red | 26 | 32.9 |
| blond | 15 | 19.0 |
| ginger | 14 | 17.7 |
| brown | 12 | 15.2 |
| auburn | 7 | 8.9 |
| other | 5 | 6.3 |

**Supplementary Table 4.** Text answers of ALSPAC young people who reported having "other" eye colour. Percentages over the total number of answers are shown.

| eye colour category | n = 257 answers | % |
|---|---|---|
| hazel | 110 | 42.8 |
| green | 56 | 21.8 |
| blue | 35 | 13.6 |
| black/brown | 27 | 10.5 |
| grey | 14 | 5.4 |
| other | 10 | 3.9 |
| changing | 5 | 1.9 |

**Supplementary Table 6.** Text answers of ALSPAC young people who reported taking “other” measures for sun protection. Percentages over the total number of answers are shown.

| sun protective measures | n = 48 answers | % |
|---|---|---|
| sit in the shade | 13 | 27.1 |
| use protection sometimes & frequent breaks from the sun & physical barriers such as clothing | 9 | 18.8 |
| sunscreen products | 8 | 16.7 |
| sunglasses | 6 | 12.5 |
| moisturizing products & tanning oils | 6 | 12.5 |
| other measures (drink water, wear make-up, depends how hot it is, all of the above, lip protection) | 6 | 12.5 |

**Supplementary Table 7.** Factors associated with reasons for liking to tan. Only those with the strongest evidence for association are shown.

| reason for tanning | it gives YP more confidence[a] | it makes YP feel happier[a] | makes YP look better in photos[a] | makes YP look thinner[a] | conceals body imperfections[a] | YP looks more attractive to others[a] | pale skin is unattractive[a] |
|---|---|---|---|---|---|---|---|
| **sex** (0=female/1=male) (N=1930) | 0.34 (0.27,0.42), $< 10^{-26}$ | 0.44 (0.35,0.54), $1.35 \times 10^{-13}$ | 0.67 (0.54,0.82), $1.57 \times 10^{-4}$ | 0.05 (0.03,0.09), $< 10^{-26}$ | 0.14 (0.09,0.20), $< 10^{-26}$ | 1.17 (0.95,1.44), 0.136 | 0.81 (0.62,1.05), 0.106 |
| **mother's social class** (0=manual/1=non-manual) (N=1598) | 1.33 (0.98,1.80), 0.068 | 1.19 (0.86,1.64), 0.290 | 0.94 (0.70,1.26), 0.695 | 1.04 (0.74,1.45), 0.837 | 1.23 (0.88,1.72), 0.223 | 1.27 (0.95,1.70), 0.109 | 0.61 (0.44,0.84), 0.003 |
| **father's social class** (0=manual/1=non-manual) (N=1704) | 0.96 (0.78,1.20), 0.743 | 1.02 (0.81,1.27), 0.875 | 1.10 (0.90,1.34), 0.357 | 1.00 (0.79,1.27), 0.996 | 1.00 (0.79,1.25), 0.973 | 1.26 (1.03,1.53), 0.025 | 0.78 (0.61,0.99), 0.041 |
| **maternal education** (N=1814) (model p-value)[1] | 0.834 | 0.250 | 0.989 | 0.578 | 0.099 | 0.007 | 0.039 |
| O level | 1.09 (0.82,1.45), 0.548 | 1.19 (0.88,1.60), 0.260 | 1.02 (0.78,1.33), 0.896 | 0.99 (0.73,1.36), 0.971 | 1.33 (0.97,1.81), 0.078 | 1.33 (1.02,1.74), 0.037 | 0.77 (0.57,1.05), 0.105 |
| > O level | 1.05 (0.80,1.38), 0.717 | 0.98 (0.74,1.29), 0.866 | 1.00 (0.78,1.29), 0.972 | 0.88 (0.65,1.20), 0.423 | 1.38 (1.02,1.87), 0.034 | 1.50 (1.16,1.93), 0.002 | 0.68 (0.51,0.91), 0.011 |
| **skin colour** (N=1929) (model p-value)[2] | 0.023 | 0.038 | 0.013 | 0.386 | 0.039 | 0.004 | $4.99 \times 10^{-5}$ |
| fair | 0.64 (0.45,0.89), 0.008 | 1.31 (0.96,1.79), 0.093 | 0.74 (0.55,1.00), 0.051 | 0.85 (0.61,1.17), 0.318 | 0.73 (0.53,0.99), 0.044 | 0.66 (0.49,0.88), 0.005 | 0.55 (0.41,0.76), $< 2.31 \times 10^{-4}$ |

| | | | | | | | |
|---|---|---|---|---|---|---|---|
| olive | 0.81 (0.56,1.16), 0.249 | 1.58 (1.11,2.24), 0.011 | 0.92 (0.66,1.28), 0.633 | 0.91 (0.63,1.31), 0.622 | 0.83 (0.59,1.18), 0.297 | 0.78 (0.57,1.08), 0.130 | 0.44 (0.31,0.64), < $1.15 \times 10^{-5}$ |
| brown | 0.64 (0.36,1.14), 0.130 | 1.98 (1.07,3.68), 0.030 | 0.49 (0.28,0.84), 0.009 | 0.54 (0.25,1.14), 0.107 | 0.39 (0.18,0.82), 0.013 | 0.43 (0.25,0.74), 0.003 | 0.33 (0.16,0.68), 0.003 |
| **tanning ability** (N=1923) (model p-value)[3] | 0.023 | 0.004 | 0.035 | 0.760 | 0.080 | 0.049 | $4.78 \times 10^{-7}$ |
| burns easily rarely tans | 0.61 (0.23,1.67), 0.339 | 0.10 (0.01,0.77), 0.027 | 0.44 (0.17,1.12), 0.085 | 0.80 (0.35,1.81), 0.593 | 0.44 (0.2,0.98), 0.044 | 0.80 (0.36,1.76), 0.574 | 0.50 (0.23,1.10), 0.085 |
| doesn't change | 0.68 (0.23,2.05), 0.496 | 0.11 (0.01,0.83), 0.033 | 0.40 (0.14,1.09), 0.074 | 1.07 (0.42,2.76), 0.887 | 0.37 (0.15,0.95), 0.038 | 0.69 (0.28,1.68), 0.409 | 0.36 (0.14,0.92), 0.033 |
| tans easily rarely burns | 0.54 (0.20,1.45), 0.220 | 0.13 (0.02,0.98), 0.048 | 0.41 (0.16,1.03), 0.057 | 0.77 (0.34,1.71), 0.518 | 0.39 (0.18,0.84), 0.017 | 0.61 (0.28,1.33), 0.212 | 0.26 (0.12,0.56), $5.76 \times 10^{-4}$ |
| always tans never burns | 0.74 (0.25,2.21), 0.593 | 0.13 (0.02,1.02), 0.052 | 0.38 (0.14,1.04), 0.059 | 0.66 (0.25,1.72), 0.395 | 0.47 (0.19,1.16), 0.100 | 0.42 (0.17,1.03), 0.059 | 0.30 (0.12,0.75), 0.010 |
| can't say, skin always protected | 0.22 (0.07,0.71), 0.011 | 0.05 (0.01,0.41), 0.005 | 0.17 (0.06,0.51), 0.002 | 0.62 (0.21,1.87), 0.398 | 0.20 (0.06,0.61), 0.005 | 0.54 (0.20,1.43), 0.214 | 0.23 (0.07,0.68), 0.009 |
| **sunburning past 2 years** (N=1868) (model p-value)[4] | 0.041 | 0.190 | $1.79 \times 10^{-4}$ | 0.053 | 0.112 | $1.52 \times 10^{-6}$ | 0.012 |
| once | 1.35 (1.02,1.80), 0.037 | 0.82 (0.61,1.11), 0.196 | 1.60 (1.22,2.09), $6.24 \times 10^{-4}$ | 1.28 (0.92,1.78), 0.138 | 1.31 (0.96,1.80), 0.086 | 1.51 (1.15,1.98), 0.003 | 1.28 (0.91,1.81), 0.160 |
| twice | 1.18 (0.90,1.56), 0.235 | 0.77 (0.57,1.03), 0.073 | 1.29 (0.99,1.67), 0.059 | 1.24 (0.89,1.71), 0.200 | 1.20 (0.88,1.64), 0.247 | 1.65 (1.26,2.15), $2.37 \times 10^{-4}$ | 1.47 (1.06,2.06), 0.023 |

| | | | | | | | |
|---|---|---|---|---|---|---|---|
| 3times | 1.56 (1.08,2.26), 0.018 | 1.13 (0.76,1.68), 0.544 | 2.03 (1.43,2.88), $6.62 \times 10^{-5}$ | 1.44 (0.96,2.17), 0.077 | 1.69 (1.15,2.48), 0.007 | 2.06 (1.46,2.90), $3.37 \times 10^{-5}$ | 1.72 (1.15,2.60), 0.009 |
| 4times | 1.49 (0.96,2.29), 0.073 | 1.08 (0.68,1.70), 0.756 | 1.65 (1.10,2.47), 0.015 | 1.24 (0.76,2.03), 0.395 | 1.36 (0.85,2.16), 0.196 | 2.68 (1.78,4.03), $2.21 \times 10^{-6}$ | 1.27 (0.77,2.10), 0.349 |
| 5times or more | 1.98 (1.16,3.36), 0.012 | 0.97 (0.57,1.66), 0.924 | 2.08 (1.28,3.37), 0.003 | 2.39 (1.40,4.11), 0.002 | 1.63 (0.96,2.77), 0.069 | 1.93 (1.21,3.08), 0.006 | 2.43 (1.44,4.10), $9.19 \times 10^{-4}$ |
| **YP does not use sun protection** (N=1928) | 0.92 (0.64,1.34), 0.667 | 0.67 (0.47,0.97), 0.035 | 0.84 (0.59,1.20), 0.341 | 1.06 (0.68,1.64), 0.799 | 1.24 (0.83,1.86), 0.297 | 1.29 (0.91,1.82), 0.160 | 1.82 (1.24,2.65), 0.002 |
| **wears sunblock** (N=1928) | 1.47 (1.01,2.14), 0.046 | 1.38 (0.94,2.02), 0.103 | 1.64 (1.13,2.36), 0.008 | 1.07 (0.67,1.71), 0.764 | 1.23 (0.79,1.94), 0.362 | 0.93 (0.65,1.34), 0.690 | 0.73 (0.48,1.10), 0.132 |
| **wears a hat** (N=1928) | 0.93 (0.75,1.17), 0.550 | 1.19 (0.93,1.50), 0.161 | 0.97 (0.78,1.20), 0.768 | 0.95 (0.73,1.23), 0.689 | 0.93 (0.73,1.19), 0.572 | 0.74 (0.60,0.91), 0.005 | 0.85 (0.66,1.10), 0.225 |
| **tanning helps prevent sunburn** (N=1926) (model p-value)[5] | $1.76 \times 10^{-4}$ | 0.001 | $4.31 \times 10^{-6}$ | $8.42 \times 10^{-6}$ | 0.006 | 0.044 | $3.91 \times 10^{-5}$ |
| yes | 2.06 (1.47,2.90), $3.21 \times 10^{-5}$ | 1.95 (1.36,2.80), $2.56 \times 10^{-4}$ | 2.18 (1.60,2.98), $7.29 \times 10^{-7}$ | 2.07 (1.51,2.83), $6.31 \times 10^{-6}$ | 1.65 (1.21,2.24), 0.001 | 1.39 (1.05,1.84), 0.023 | 2.02 (1.49,2.74), $6.95 \times 10^{-6}$ |
| don’t know | 1.08 (0.84,1.39), 0.529 | 1.13 (0.87,1.46), 0.373 | 1.05 (0.83,1.33), 0.694 | 0.87 (0.65,1.18), 0.381 | 1.13 (0.85,1.49), 0.369 | 0.93 (0.74,1.17), 0.532 | 1.09 (0.82,1.46), 0.534 |
| **indoor tanning can cause skin cancer** (N=1924) (model p-value)[5] | 0.005 | 0.013 | 0.348 | $2.36 \times 10^{-4}$ | 0.005 | 0.001 | 0.107 |
| yes | 1.16 (0.64,2.10), 0.631 | 1.45 (0.81,2.62), 0.213 | 0.98 (0.55,1.72), 0.931 | 0.64 (0.35,1.18), 0.155 | 0.90 (0.49,1.65), 0.732 | 1.03 (0.59,1.79), 0.921 | 0.61 (0.34,1.10), 0.101 |

| | | | | | | | |
|---|---|---|---|---|---|---|---|
| don't know | 0.72 (0.37,1.39), 0.326 | 1.26 (0.65,2.45), 0.489 | 0.58 (0.31,1.09), 0.091 | 0.25 (0.12,0.54), $3.88x10^{-4}$ | 0.45 (0.21,0.92), 0.030 | 0.54 (0.29,1.02), 0.057 | 0.47 (0.24,0.95), 0.035 |
| **YP trying to change skin colour** (N=1537) | 2.36 (1.54,3.62), $8.55x10^{-5}$ | 1.62 (1.07,2.47), 0.023 | 1.92 (1.34,2.76), $3.75x10^{-4}$ | 2.06 (1.45,2.94), $5.95x10^{-5}$ | 1.69 (1.19,2.39), 0.003 | 1.73 (1.24,2.42), 0.001 | 1.79 (1.25,2.56), 0.001 |
| **YP trying to change hair colour** (N=1537) | 1.48 (0.94,2.35), 0.094 | 1.53 (0.93,2.51), 0.091 | 2.14 (1.38,3.31), $7.05x10^{-4}$ | 1.99 (1.32,3.00), 0.001 | 2.09 (1.39,3.13), $3.52x10^{-4}$ | 1.25 (0.85,1.85), 0.264 | 1.41 (0.91,2.19), 0.123 |
| **YP's teachers treat everybody the same** (N=1478) | 1.51 (0.82,2.78), 0.189 | 1.41 (0.74,2.68), 0.300 | 0.90 (0.52,1.54), 0.695 | 1.62 (0.85,3.10), 0.146 | 1.44 (0.77,2.68), 0.253 | 0.63 (0.36,1.10), 0.104 | 2.04 (1.15,3.63), 0.015 |

[a]OR (95% CI), p-value, from logistic regression model adjusted for age and sex.

[1]reference: < O level

[2]reference: very fair

[3]reference: always burns, never tans

[4]reference: never

[5]reference: no

**Supplementary Table 8.** Association of socio-demographic, pigmentation-related and sun exposure factors with outdoor tanning in ALSPAC young people who like to tan.

| exposure | OR[a] | 95% CI | p-value | N |
|---|---|---|---|---|
| **sex** (0=female/1=male) | 6.11 | (3.29,11.32) | $9.35 \times 10^{-9}$ | 1929 |
| **age** (years) | 1.25 | (0.96,1.62) | 0.103 | 1929 |
| **mother's social class** (0=manual/1=non-manual) | 1.66 | (1.09,2.52) | 0.018 | 1597 |
| **father's social class** (0=manual/1=non-manual) | 1.60 | (1.15,2.21) | 0.005 | 1703 |
| **mother's education** | | | $3.83 \times 10^{-5}$ | 1813 |
| < O level | reference | | | |
| O level | 1.68 | (1.14,2.47) | 0.009 | |
| > O level | 2.44 | (1.66,3.61) | $6.55 \times 10^{-6}$ | |
| **skin colour** | | | $2.71 \times 10^{-12}$ | 1928 |
| very fair | reference | | | |
| fair | 3.09 | (2.14,4.44) | $1.35 \times 10^{-9}$ | |
| olive | 5.30 | (3.25,8.66) | $2.66 \times 10^{-11}$ | |
| brown | 9.42 | (2.22,40.03) | 0.002 | |
| **eye colour** | | | 0.942 | 1925 |
| blue | reference | | | |
| green | 1.07 | (0.72,1.58) | 0.748 | |
| grey | 1.56 | (0.55,4.47) | 0.405 | |
| brown | 1.06 | (0.74,1.52) | 0.766 | |
| other | 0.99 | (0.49,2.00) | 0.980 | |
| **natural hair colour at 18 years old** | | | $8.39 \times 10^{-6}$ | 1908 |
| red | reference | | | |
| blond | 3.72 | (1.85,7.44) | $2.14 \times 10^{-4}$ | |
| light brown | 5.10 | (2.60,9.98) | $2.02 \times 10^{-6}$ | |
| dark brown | 5.94 | (3.00,11.80) | $3.43 \times 10^{-7}$ | |

| | | | | |
|---|---|---|---|---|
| black | 1.00 | n/a | n/a | |
| other | 3.32 | (0.96,11.50) | 0.058 | |
| **YP has freckles** | | | 0.004 | 1925 |
| no | reference | | | |
| yes, a few | 0.62 | (0.44,0.87) | 0.006 | |
| yes, many | 0.51 | (0.32,0.80) | 0.003 | |
| **tanning ability** | | | $1.69x10^{-17}$ | 1922 |
| always burns never tans | reference | | | |
| burns easily rarely tans | 8.24 | (3.50,19.41) | $1.37x10^{-6}$ | |
| doesn't change | 4.69 | (1.75,12.55) | 0.002 | |
| tans easily rarely burns | 23.79 | (10.13,55.86) | $3.41x10^{-13}$ | |
| always tans never burns | 14.21 | (4.72,42.79) | $2.40x10^{-6}$ | |
| can't say, skin always protected | 14.78 | (3.93,55.58) | $6.72x10^{-5}$ | |
| **sun burning in the past 2 years** | | | 0.022 | 1867 |
| never | reference | | | |
| once | 1.32 | (0.88,1.98) | 0.178 | |
| twice | 1.93 | (1.25,2.97) | 0.003 | |
| 3 times | 1.89 | (1.06,3.38) | 0.031 | |
| 4 times | 1.57 | (0.80,3.07) | 0.186 | |
| 5 times or more | 0.84 | (0.43,1.64) | 0.611 | |
| **sun protection** (0=no/1=yes) | | | | |
| never uses protection | 0.70 | (0.41,1.20) | 0.193 | 1927 |
| wears hat | 1.21 | (0.83,1.75) | 0.317 | 1927 |
| wears clothing | 0.89 | (0.63,1.25) | 0.496 | 1927 |
| wears sunblock | 1.35 | (0.76,2.40) | 0.298 | 1927 |
| avoids the sun | 0.83 | (0.50,1.38) | 0.463 | 1927 |
| **sunblock factor** | | | 0.162 | 1797 |
| lower than 15 | reference | | | |

| | | | | |
|---|---|---|---|---|
| 15-24 | 0.94 | (0.49,1.80) | 0.856 | |
| 25-49 | 0.80 | (0.42,1.52) | 0.492 | |
| 50 or higher | 0.52 | (0.24,1.11) | 0.092 | |
| **skin cancer** (0=no/1=yes) | | | | |
| YP has family member diagnosed with skin cancer | 0.82 | (0.54,1.24) | 0.339 | 1924 |
| **beliefs about tanning** | | | | |
| YP believes indoor tanning helps prevent sunburn | | | 0.016 | 1925 |
| no | reference | | | |
| yes | 0.55 | (0.37,0.83) | 0.004 | |
| don't know | 0.86 | (0.58,1.28) | 0.452 | |
| YP thinks indoor tanning using sun bed/sun lamp can cause skin cancer | | | 0.580 | 1923 |
| no | reference | | | |
| yes | 1.51 | (0.69,3.30) | 0.300 | |
| don't know | 1.44 | (0.57,3.60) | 0.437 | |
| **body image** | | | | |
| YP is trying to change skin colour (0=no/1=yes) | 0.84 | (0.50,1.39) | 0.490 | 1536 |
| YP is trying to change hair colour (0=no/1=yes) | 0.60 | (0.35,1.03) | 0.063 | 1536 |
| YP has been hurt because of their skin colour (0=no/1=yes) | 0.66 | (0.08,5.42) | 0.695 | 1603 |
| YP has been called names because of their skin colour (0=no/1=yes) | 0.50 | (0.18,1.36) | 0.175 | 1607 |
| YP has seen someone bullied because of their skin colour (0=no/1=yes) | 1.07 | (0.73,1.57) | 0.741 | 1598 |
| YP's teachers treat everybody the same, regardless of skin colour (0=agree/1=disagree) | 0.70 | (0.29,1.71) | 0.438 | 1477 |

[a]Adjusted for age and sex.

**Supplementary Table 9.** Association of socio-demographic, pigmentation-related and sun exposure factors with indoor tanning in the past 12 months amongst ALSPAC young people who manifested ever engaging in indoor tanning using a sun bed/sun lamp/tanning booth.

| exposure | OR[a] | 95% CI | p-value | N |
|---|---|---|---|---|
| **sex** (0=female/1=male) | 0.79 | (0.54,1.15) | 0.216 | 946 |
| **age** (years) | 0.83 | (0.66,1.03) | 0.095 | 946 |
| **mother's social class** (0=manual/1=non-manual) | 1.01 | (0.69,1.47) | 0.970 | 775 |
| **father's social class** (0=manual/1=non-manual) | 0.61 | (0.46,0.80) | $4.46 \times 10^{-4}$ | 837 |
| **mother's education** | | | 0.014 | 884 |
| < O level | reference | | | |
| O level | 1.18 | (0.83,1.67) | 0.356 | |
| > O level | 0.75 | (0.53,1.06) | 0.108 | |
| **skin colour** | | | 0.003 | 943 |
| very fair | reference | | | |
| fair | 1.60 | (1.08,2.38) | 0.019 | |
| olive | 2.24 | (1.44,3.50) | $3.57 \times 10^{-4}$ | |
| brown | 2.60 | (1.03,6.55) | 0.043 | |
| **eye colour** | | | 0.131 | 945 |
| blue | reference | | | |
| green | 1.47 | (1.05,2.06) | 0.025 | |
| grey | 2.06 | (0.88,4.82) | 0.096 | |
| brown | 1.21 | (0.89,1.65) | 0.227 | |
| other | 1.08 | (0.56,2.09) | 0.823 | |
| **natural hair colour at 18 years old** | | | 0.725 | 946 |
| red | reference | | | |
| blonde | 1.97 | (0.65,6.00) | 0.231 | |
| light brown | 1.95 | (0.65,5.81) | 0.233 | |

| | | | | |
|---|---|---|---|---|
| dark brown | 2.00 | (0.67,6.01) | 0.215 | |
| black | 0.60 | (0.05,7.03) | 0.681 | |
| other | 2.62 | (0.48,14.35) | 0.267 | |
| **YP has freckles** | | | 0.044 | 944 |
| no | reference | | | |
| yes, a few | 1.15 | (0.87,1.51) | 0.333 | |
| yes, many | 0.69 | (0.46,1.04) | 0.076 | |
| **tanning ability** | | | $1.65 \times 10^{-4}$ | 939 |
| always burns never tans | reference | | | |
| burns easily rarely tans | 2.25 | (0.97,5.22) | 0.060 | |
| doesn't change | 1.83 | (0.64,5.26) | 0.260 | |
| tans easily rarely burns | 3.51 | (1.55,7.99) | 0.003 | |
| always tans never burns | 7.11 | (2.55,19.81) | $1.73 \times 10^{-4}$ | |
| can’t say, skin always protected | 2.62 | (0.69,9.98) | 0.159 | |
| **sun burning in the past 2 years** | | | 0.132 | 906 |
| never | reference | | | |
| once | 0.71 | (0.49,1.02) | 0.066 | |
| twice | 0.63 | (0.43,0.92) | 0.017 | |
| 3 times | 0.72 | (0.44,1.17) | 0.182 | |
| 4 times | 1.07 | (0.59,1.93) | 0.832 | |
| 5 times or more | 0.99 | (0.53,1.83) | 0.964 | |
| **sun protection** (0=no/1=yes) | | | | |
| never uses protection | 1.28 | (0.77,2.11) | 0.337 | 944 |
| wears hat | 0.83 | (0.62,1.13) | 0.244 | 944 |
| wears clothing | 0.68 | (0.50,0.94) | 0.019 | 944 |
| wears sunblock | 0.79 | (0.47,1.34) | 0.390 | 944 |
| avoids the sun | 0.55 | (0.33,0.91) | 0.020 | 944 |
| **sunblock factor** | | | $1.18 \times 10^{-7}$ | 883 |

| | | | | |
|---|---|---|---|---|
| lower than 15 | reference | | | |
| 15-24 | 0.75 | (0.47,1.20) | 0.231 | |
| 25-49 | 0.42 | (0.26,0.67) | $2.72 \times 10^{-4}$ | |
| 50 or higher | 0.20 | (0.10,0.41) | $8.39 \times 10^{-6}$ | |
| **skin cancer** (0=no/1=yes) | | | | |
| YP has family member diagnosed with skin cancer | 1.13 | (0.76,1.67) | 0.548 | 944 |
| **beliefs about tanning** (0=no/1=yes) | | | | |
| YP believes indoor tanning helps prevent sunburn | | | $9.04 \times 10^{-14}$ | 946 |
| no | reference | | | |
| yes | 4.10 | (2.87,5.86) | $1.04 \times 10^{-14}$ | |
| don't know | 1.25 | (0.88,1.76) | 0.213 | |
| YP thinks indoor tanning using sun bed/sun lamp can cause skin cancer | | | 0.003 | 943 |
| no | reference | | | |
| yes | 0.36 | (0.16,0.83) | 0.016 | |
| don't know | 0.70 | (0.27,1.84) | 0.472 | |
| **body image** | | | | |
| YP is trying to change skin colour (0=no/1=yes) | 0.90 | (0.60,1.35) | 0.615 | 731 |
| YP is trying to change hair colour (0=no/1=yes) | 0.89 | (0.55,1.43) | 0.627 | 731 |
| YP has been hurt because of their skin colour (0=no/1=yes) | 0.74 | (0.12,4.55) | 0.750 | 774 |
| YP has been called names because of their skin colour (0=no/1=yes) | 1.08 | (0.34,3.38) | 0.899 | 780 |
| YP has seen someone bullied because of their skin colour (0=no/1=yes) | 0.96 | (0.70,1.30) | 0.777 | 774 |
| YP's teachers treat everybody the same, regardless of skin colour (0=agree/1=disagree) | 0.76 | (0.35,1.62) | 0.473 | 684 |

[a]Adjusted for age and sex.

**Supplementary Table 10.** Association of socio-demographic, pigmentation-related and sun exposure factors with indoor tanning using spray tan or self-tanning lotions amongst ALSPAC young people who like to tan.

| exposure | OR[a] | 95% CI | p-value | N |
|---|---|---|---|---|
| **sex** (0=female/1=male) | 0.05 | (0.03,0.09) | < $10x^{-26}$ | 1930 |
| **age** (years) | 0.99 | (0.82,1.19) | 0.908 | 1930 |
| **mother's social class** (0=manual/1=non-manual) | 0.87 | (0.63,1.20) | 0.401 | 1598 |
| **father's social class** (0=manual/1=non-manual) | 0.73 | (0.58,0.91) | 0.006 | 1704 |
| **mother's education** | | | 0.004 | 1814 |
| < O level | reference | | | |
| O level | 0.75 | (0.56,1.01) | 0.060 | |
| > O level | 0.62 | (0.46,0.82) | 0.001 | |
| **skin colour** | | | $6.23x10^{-7}$ | 1929 |
| very fair | reference | | | |
| fair | 0.60 | (0.44,0.82) | 0.001 | |
| olive | 0.38 | (0.26,0.54) | $7.94x10^{-8}$ | |
| brown | 0.35 | (0.17,0.71) | 0.004 | |
| **eye colour** | | | 0.348 | 1926 |
| blue | reference | | | |
| green | 0.88 | (0.67,1.16) | 0.363 | |
| grey | 0.92 | (0.50,1.72) | 0.804 | |
| brown | 0.79 | (0.62,1.03) | 0.077 | |
| other | 1.18 | (0.73,1.92) | 0.495 | |
| **natural hair colour at 18 years old** | | | 0.045 | 1927 |
| red | reference | | | |
| blonde | 0.68 | (0.35,1.33) | 0.260 | |

| | | | | |
|---|---|---|---|---|
| light brown | 0.48 | (0.25,0.92) | 0.027 | |
| dark brown | 0.51 | (0.26,0.98) | 0.042 | |
| black | 0.96 | (0.11,8.48) | 0.974 | |
| other | 0.75 | (0.27,2.08) | 0.587 | |
| **YP has freckles** | | | 0.006 | 1926 |
| no | reference | | | |
| yes, a few | 1.41 | (1.12,1.77) | 0.003 | |
| yes, many | 1.47 | (1.06,2.04) | 0.019 | |
| **tanning ability** | | | $5.10 \times 10^{-9}$ | 1923 |
| always burns never tans | reference | | | |
| burns easily rarely tans | 0.16 | (0.06,0.46) | 0.001 | |
| doesn't change | 0.21 | (0.07,0.66) | 0.008 | |
| tans easily rarely burns | 0.09 | (0.03,0.26) | $5.28 \times 10^{-6}$ | |
| always tans never burns | 0.12 | (0.04,0.37) | $2.41 \times 10^{-4}$ | |
| **sun burning in the past 2 years** | | | 0.290 | 1868 |
| never | reference | | | |
| once | 0.89 | (0.66,1.20) | 0.443 | |
| twice | 0.78 | (0.58,1.06) | 0.112 | |
| 3 times | 0.86 | (0.58,1.26) | 0.432 | |
| 4 times | 1.17 | (0.74,1.85) | 0.495 | |
| 5 times or more | 1.23 | (0.73,2.10) | 0.436 | |
| **sun protection** (0=no/1=yes) | | | | |
| never uses protection | 0.78 | (0.51,1.22) | 0.277 | 1928 |
| wears hat | 1.12 | (0.87,1.43) | 0.377 | 1928 |
| wears clothing | 0.97 | (0.76,1.24) | 0.791 | 1928 |
| wears sunblock | 1.73 | (1.07,2.81) | 0.027 | 1928 |
| avoids the sun | 1.03 | (0.71,1.50) | 0.872 | 1928 |
| **sunblock factor** | | | 0.074 | 1798 |

| | | | | |
|---|---|---|---|---|
| lower than 15 | reference | | | |
| 15-24 | 0.63 | (0.42,0.95) | 0.027 | |
| 25-49 | 0.58 | (0.39,0.87) | 0.008 | |
| 50 or higher | 0.62 | (0.37,1.07) | 0.086 | |
| **skin cancer** (0=no/1=yes) | | | | |
| YP has family member diagnosed with skin cancer | 1.11 | (0.82,1.50) | 0.492 | 1925 |
| **beliefs about tanning** (0=no/1=yes) | | | | |
| YP believes indoor tanning helps prevent sunburn | | | $2.57 \times 10^{-4}$ | 1926 |
| no | reference | | | |
| yes | 1.84 | (1.35,2.51) | $1.19 \times 10^{-4}$ | |
| don’t know | 0.92 | (0.69,1.22) | 0.544 | |
| YP thinks indoor tanning using sun bed/sun lamp can cause skin cancer | | | 0.021 | 1924 |
| no | reference | | | |
| yes | 1.18 | (0.62,2.22) | 0.617 | |
| don’t know | 0.66 | (0.31,1.38) | 0.266 | |
| **body image** | | | | |
| YP is trying to change skin colour (0=no/1=yes) | 1.71 | (1.20,2.42) | 0.003 | 1537 |
| YP is trying to change hair colour (0=no/1=yes) | 2.10 | (1.39,3.15) | $3.70 \times 10^{-4}$ | 1537 |
| YP has been hurt because of their skin colour (0=no/1=yes) | 1.54 | (0.33,7.16) | 0.508 | 1604 |
| YP has been called names because of their skin colour (0=no/1=yes) | 1.34 | (0.58,3.08) | 0.495 | 1608 |
| YP has seen someone bullied because of their skin colour (0=no/1=yes) | 1.11 | (0.86,1.44) | 0.414 | 1599 |
| YP’s teachers treat everybody the same, regardless of skin colour (0=agree/1=disagree) | 2.30 | (1.21,4.37) | 0.011 | 1478 |

[a]Adjusted for age and sex.

**Supplementary Table 16.** Association of indoor tanning using sprays or self-tanning lotions with HIrisPlex-S prediction probabilities of pigmentation traits in ALSPAC young people (N = 1489).

| HIrisPlex-S probabilities | OR[a] | 95% | p-value |
|---|---|---|---|
| PBlueEye | 1.12 | (0.82,1.52) | 0.477 |
| PIntermediateEye | 1.14 | (0.22,5.99) | 0.879 |
| PBrownEye | 0.87 | (0.62,1.22) | 0.415 |
| PBlondHair | 0.86 | (0.51,1.45) | 0.580 |
| PBrownHair | 0.97 | (0.51,1.84) | 0.932 |
| PRedHair | 2.15 | (0.95,4.85) | 0.064 |
| PBlackHair | 0.36 | (0.09,1.49) | 0.161 |
| PLightHair | 1.41 | (0.86,2.31) | 0.176 |
| PDarkHair | 0.71 | (0.43,1.17) | 0.176 |
| PVeryPaleSkin | 5.16 | (1.31,20.32) | 0.019 |
| PPaleSkin | 1.49 | (0.73,3.05) | 0.277 |
| PIntermediateSkin | 0.57 | (0.31,1.07) | 0.078 |
| PDarkSkin | 0.03 | ($2.37 \times 10^{-4}$,4.10) | 0.164 |
| PDarktoBlackSkin | 0.06 | ($5.66 \times 10^{-9}$,$6.16 \times 10^{5}$) | 0.731 |

[a]Adjusted for age, sex and 10 PCs.
